# Diagnostic Accuracy of Commercially Available Tests for Respiratory Syncytial Virus: A Scoping Literature Review in the COVID-19 Era

**DOI:** 10.1101/2022.02.14.22270927

**Authors:** David I. Bernstein, Asuncion Mejias, Barbara Rath, Christopher W. Woods, Jamie Phillips Deeter

## Abstract

**Background:** Non-pharmaceutical interventions to prevent the spread of coronavirus disease 2019 also decreased the spread of respiratory syncytial virus (RSV) and influenza. Viral diagnostic testing in patients with respiratory tract infections (RTI) is a necessary tool for patient management; therefore, sensitive and specific tests are required. This scoping literature review evaluated the analytical validity of commercially available sample-to- answer RSV diagnostic tests in different contexts.

**Content:** PubMed and Embase were queried for studies reporting on the analytical validity of tests for RSV in patients with RTI (published January 2005–January 2021). Sensitivity and specificity of RSV tests and information on study design, patient, and setting characteristics were extracted from 77 studies that met predefined inclusion criteria. A literature gap was identified for studies of RSV tests conducted in adult-only populations (5.3% of total sub- records), and in outpatient (7.5%) or household (0.8%) settings. Overall, RSV tests with analytical time >30 min had higher sensitivity (62.5–100%) versus RSV tests with analytical time ≤ 30 min (25.7–100%), this sensitivity range could be partially attributed to the different modalities (antigen versus molecular) used. Molecular-based rapid RSV tests had higher sensitivity (66.7–100%) and specificity (94.3–100%) than antigen-based RSV tests (25.7– 100%; 80.3–100%).

**Summary:** Molecular-based RSV tests should be considered for first-line use when possible, given their high sensitivity and specificity and that adults with RTI typically have low viral load, necessitating a highly sensitive test. This review benefits healthcare professionals by summarizing the diagnostic accuracy data available for commercially available RSV tests.

**IMPACT STATEMENT:** Viral diagnostic testing in patients with respiratory tract infection is a powerful tool for patient management. This scoping literature review included 77 studies reporting the analytical validity of commercially available respiratory syncytial virus (RSV) diagnostic tests (published January 2005–January 2021) and examined the characteristics of such studies. The data suggest that molecular-based RSV tests have higher sensitivity and specificity than antigen-based tests, thus should be considered for first-line use for timely diagnosis and to detect infections in adults with low level viral load. Future studies should investigate the diagnostic accuracy of RSV tests in adults and in outpatient/household settings.

## INTRODUCTION

Respiratory syncytial virus (RSV) infection is responsible for a significant proportion of outpatient visits and hospitalizations in children <5 years old and is associated with substantial clinical and economic burden (1). In a recent international study across 72 countries, there was an annual mean of 20.8 million cases and 1.8 million hospital admissions for RSV infection among children <5 years old, and this was associated with $611 million (USD) discounted direct costs (2). Once considered to be a disease of childhood, there is increasing recognition of the prevalence of RSV infection in the community dwelling (3) and hospitalized adult populations; for example, the rate of hospitalization and economic burden of hospitalized adults and the elderly (≥ 60 years old) with RSV infection has been shown to rival that of influenza in the United States (US) (4–6).

The clinical manifestations of RSV infection vary according to the age of the patient and host susceptibility to the virus, ranging from cold symptoms to severe respiratory distress (7). Both adult and pediatric patients infected with RSV often present with non-specific, overlapping symptoms that can lead to difficulty in distinguishing it from influenza, coronavirus disease 2019 (COVID-19) caused by severe acute respiratory syndrome coronavirus 2 (SARS-CoV-2), or other respiratory illnesses (8). Thus, empiric diagnosis is often insufficient and should be supported by viral diagnostic testing to facilitate appropriate treatment, improved surveillance, and timely infection control (9).

Historically, viral culture was the gold standard technique to diagnose a productive RSV infection; however, it does not provide timely results to inform clinical management (10). Therefore, real-time reverse transcription-polymerase chain reaction (rRT-PCR), which detects the presence of the virus (active or inactive) with equal or greater sensitivity than viral culture, is often referred to as the reference/gold standard for RSV diagnosis in clinical laboratories (10). There are further modalities available for the detection of RSV with variable diagnostic accuracy e.g., antigen-based testing is sensitive for detecting RSV in young children but is not sensitive enough for use in older children or adults, as per the Centers for Disease Control and Prevention (CDC) guidance, due to lower viral loads in the respiratory specimens of this group (11). Thus, rRT-PCR testing is recommended for adults with suspected RSV infection (11).

Most RSV testing takes place in hospitalized patients (12) where selection bias exists towards more severe cases and pediatric patients, the age group most likely to be hospitalized due to RSV infection (13). Testing for RSV in adults by internists and general practitioners is rare, partially due to lack of awareness (14). A recent international study conducted across 15 countries reported that cases of RSV in adults ≥ 65 years old were notably under-represented in national surveillance programs (15). RSV testing is also limited in the younger population as shown by a prospective study in pediatric patients (≤18 years old) in Germany, which revealed that only 8.7% of patients presenting with symptoms of a respiratory tract infection underwent viral diagnostic testing during standard-of-care procedures (16). The lack of routine testing for RSV may contribute to the underestimation of disease prevalence and this has practical implications. In one study based in the emergency department of a US hospital, patients aged 6–21 years old accounted for 8.7% of the total number of RSV positive tests, whereas patients aged 22–59 years old and those aged ≥ 60 years old accounted for 14.0% and 10.5%, respectively (17). Viral diagnostic testing in pediatric and adult populations helps tailor patient management and the implementation of hospital infection prevention policies, as well as reduce the inappropriate use of antibiotics (18).

Non-pharmaceutical interventions implemented to prevent the spread of SARS-CoV-2 have also impacted the spread of RSV and influenza virus, resulting in a larger population of potential immune-naïve populations, which could lead to an increase in disease burden for future respiratory virus seasons. As a result, models predict sporadic outbreaks and an increase in the prevalence of these diseases (19). Indeed, the CDC recently released a health advisory notice warning of increased interseasonal RSV activity across the Southern US (20), and a similar interseasonal resurgence of RSV has been reported in pediatric populations in another area of the US (21), Switzerland (22), and Australia (23). Viral diagnostic testing in patients presenting with symptoms of a respiratory tract infection is a powerful tool for surveillance and patient management during such periods of interseasonal resurgence and for future respiratory virus seasons.

Sample-to-answer diagnostic tests encompass technology where sample extraction, processing, and analysis are automated, and the test is often performed directly in one cartridge or analyzer. These tests are often in the form of two different modalities, a lateral flow antigen capture test or a point-of-care (POC) molecular test. Such tests offer key advantages over laboratory-based diagnostics including ease of use and faster turnaround time (24). Reducing the time needed to diagnose RSV has been shown to be beneficial in adult and pediatric populations. The duration of time to RSV diagnosis from the point of hospital admission to test result is positively correlated with length of hospital stay and antibiotic use in hospitalized adults (12). Additionally, the use of POC testing for RSV in pediatric patients has been associated with a reduction in: the use of antibiotic treatment, the need for further clinical investigations, and time spent in the emergency department (25).

The Clinical Laboratory Improvement Amendments (CLIA) include federal standards that regulate US clinical laboratories performing diagnostic testing on human samples (26). As defined by CLIA, waived tests are simple tests with a low risk for an incorrect result; non- waived testing is used to refer to moderate or high complexity tests that can only be used at clinical laboratories that meet certain quality standards (27). There are several CLIA-waived and non-waived tests available for the diagnosis of RSV. Information on the sensitivity and specificity of such tests, and the patient and setting characteristics where these tests have been studied, will help healthcare professionals’ decision-making on the most appropriate test to use in their practice.

The objective of this scoping literature review was to evaluate the sensitivity and specificity of commercially available sample-to-answer diagnostic tests for RSV in patients with acute respiratory infection and examine the characteristics of those studies. This review also sought to identify knowledge gaps in the patient and setting characteristics where these tests have been studied.

## MATERIALS AND METHODS

### Scoping Review Design, Data Sources, and Search Strategy

This scoping literature review was conducted using the Scoping Review methodology as described by Arksey and O’Malley (28) and the guidance provided in the Preferred Reporting Items for Systematic Reviews and Meta-Analyses extension for Scoping Reviews (29).

PubMed (https://pubmed.ncbi.nlm.nih.gov/) and Embase (https://www.embase.com/) were interrogated on January 21, 2021 using the search terms and criteria in Supplemental Table 1 to identify studies reporting on the sensitivity and specificity of commercially available sample-to-answer tests for RSV in patients with acute respiratory infection published between January 2005 and January 2021. Database searches were supplemented by manual searches and references, as appropriate. Duplicate articles and ineligible publication types (narrative reviews, editorials, case reports, addresses, biographies, comments, directories, Festschrifts, interviews, lectures, legal cases, legislation, news, newspaper article, patient education handouts, popular works) were excluded. The titles and abstracts of the remaining articles were reviewed by two independent reviewers in parallel and discrepancies were resolved through discussion. Full-text articles were then obtained, and a second round of screening was conducted by two reviewers working in parallel, with adjudication through discussion. Articles not meeting the inclusion criteria were excluded as necessary.

### Study Selection

This scoping literature review included any peer-reviewed studies in the English language providing original data on the sensitivity and specificity of a commercially available sample-to-answer test for RSV (using any molecular or non-molecular diagnostic tools) relative to an in-house or commercial rRT-PCR, viral culture, and/or immunofluorescence assay as the reference standard in patients of any age with symptoms of a respiratory tract infection in any setting. Original research articles, systematic reviews, and meta-analyses were included in the review. Studies that used a non-commercial RSV test, studies where the RSV and reference test were not carried out in the same samples, studies in immunocompromised patients, or studies not otherwise meeting the inclusion criteria were excluded. Health economic analyses and pre-clinical research studies were also excluded.

### Data Extraction

The following information was extracted, where available: RSV test sensitivity, RSV test specificity, commercial brand of RSV test, data collection (prospective vs retrospective), industry sponsorship, age group of study population (adults were defined as patients ≥18 years old), majority (>75%) specimen type, majority (>75%) setting of patient recruitment, and setting where the RSV test was performed. Any missing data were recorded as ‘not reported’ and included in the data synthesis. The analytical time for each test was taken from its respective manufacturer’s data sheet. Rapid tests, including those that were suitable for use at the POC, were defined as having an analytical time ≤30 min. Where one article reported several relevant sensitivity and specificity values (e.g., when more than one RSV test was studied or if there was a prospective and retrospective arm of the study), then each test or study arm was extracted as a ‘sub-record’. Following data extraction, analysis of discordant results between the two reviewers was conducted by a third independent party and discrepancies resolved.

### Data Reporting

All data handling was carried out using Microsoft Excel 365 (Microsoft Corporation). The range of sensitivity and specificity values and summary statistics were recorded including the lowest and highest value quoted for a particular RSV test from the relevant sub-records.

Sensitivity and specificity ranges were not reported for RSV tests with <3 supporting sub-records.

## RESULTS

### Literature Search Outcome

Following screening of titles and abstracts, 200 articles were subject to full-text screening and 77 studies were eventually included in the qualitative synthesis (Fig. 1). In studies reporting several relevant sensitivity and specificity values (e.g., when more than one RSV test was studied), each test was extracted as a sub-record. The 77 included studies corresponded to 133 included sub-records. Overall, the literature search detected 39 different commercially available RSV tests from 27 manufacturers, which represented a variety of technologies and analytical times (Supplemental Table 2).

**Fig. 1.**
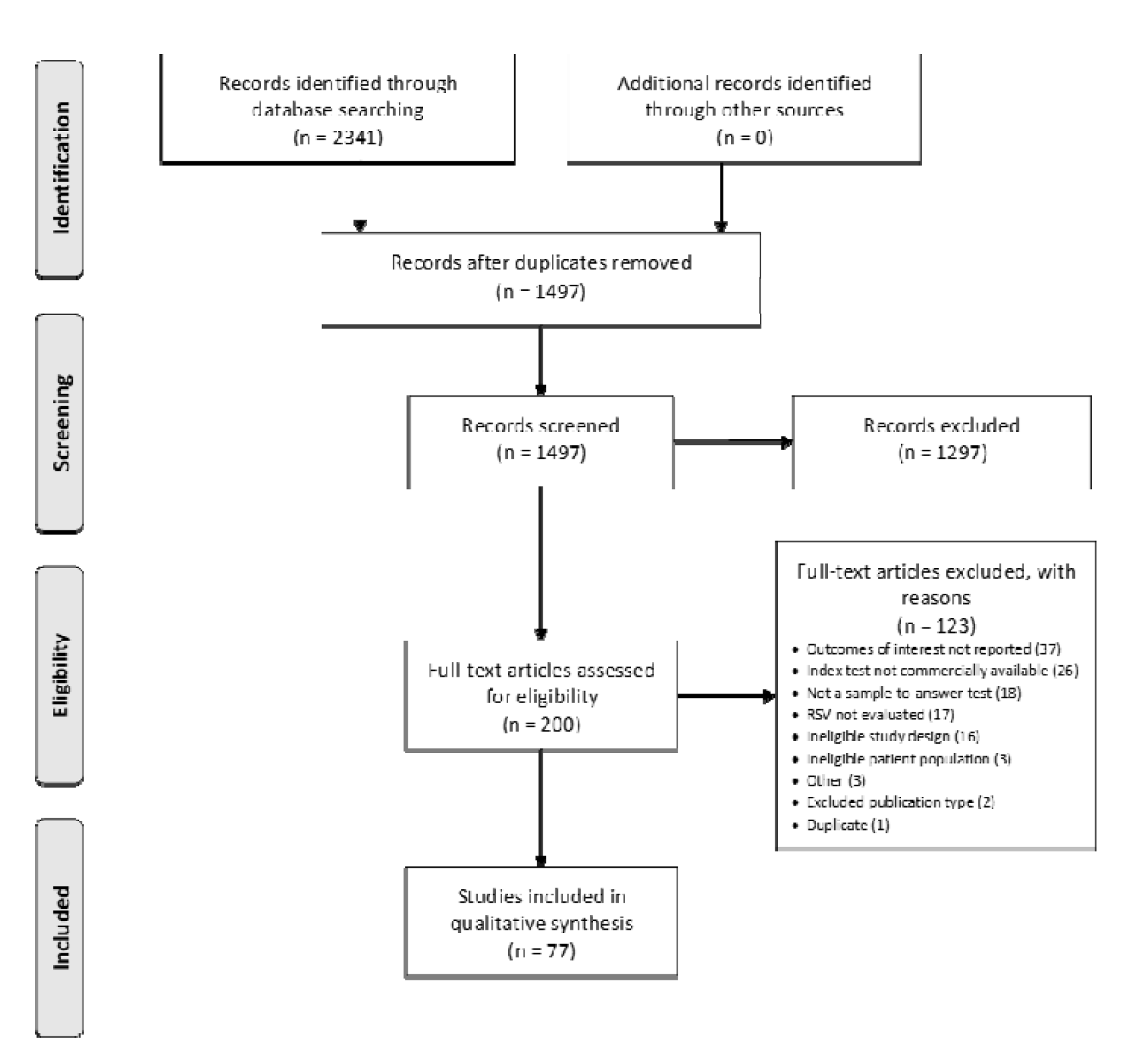
Flow chart summarizing data sources and study selection. Duplicate articles and ineligible publication types were excluded at the ‘Screening’ step (n=1297). Where one article reported several relevant sensitivity and specificity values (e.g., when more than one RSV test was studied), then each test was extracted as a sub-record. The 77 included studies corresponded to 133 included sub-records. RSV, respiratory syncytial virus.

### Characteristics of All Included Studies and Gap Analysis

Most studies examined RSV tests with analytical time ≤30 min relative to RSV tests with analytical time >30 min (66.2% vs 33.8% of included sub-records; Table 1). The analytical times taken from the manufacturer’s data sheet for each test are shown in Supplemental Table 2. Most studies assessed RSV tests in mixed (49.6% of included sub-records) or pediatric (38.3%) populations, were prospective in design (62.4% of included sub-records), used rRT-PCR as the reference standard (68.4%), and were industry sponsored (65.4%; Fig. 2). In all studies evaluated, a nasopharyngeal swab was the specimen type most used (48.9% of included sub-records; Fig. 2). Most patients were recruited when they were admitted to the hospital (42.1%) or from mixed (34.6%) settings (Fig. 2).

**Table 1.**
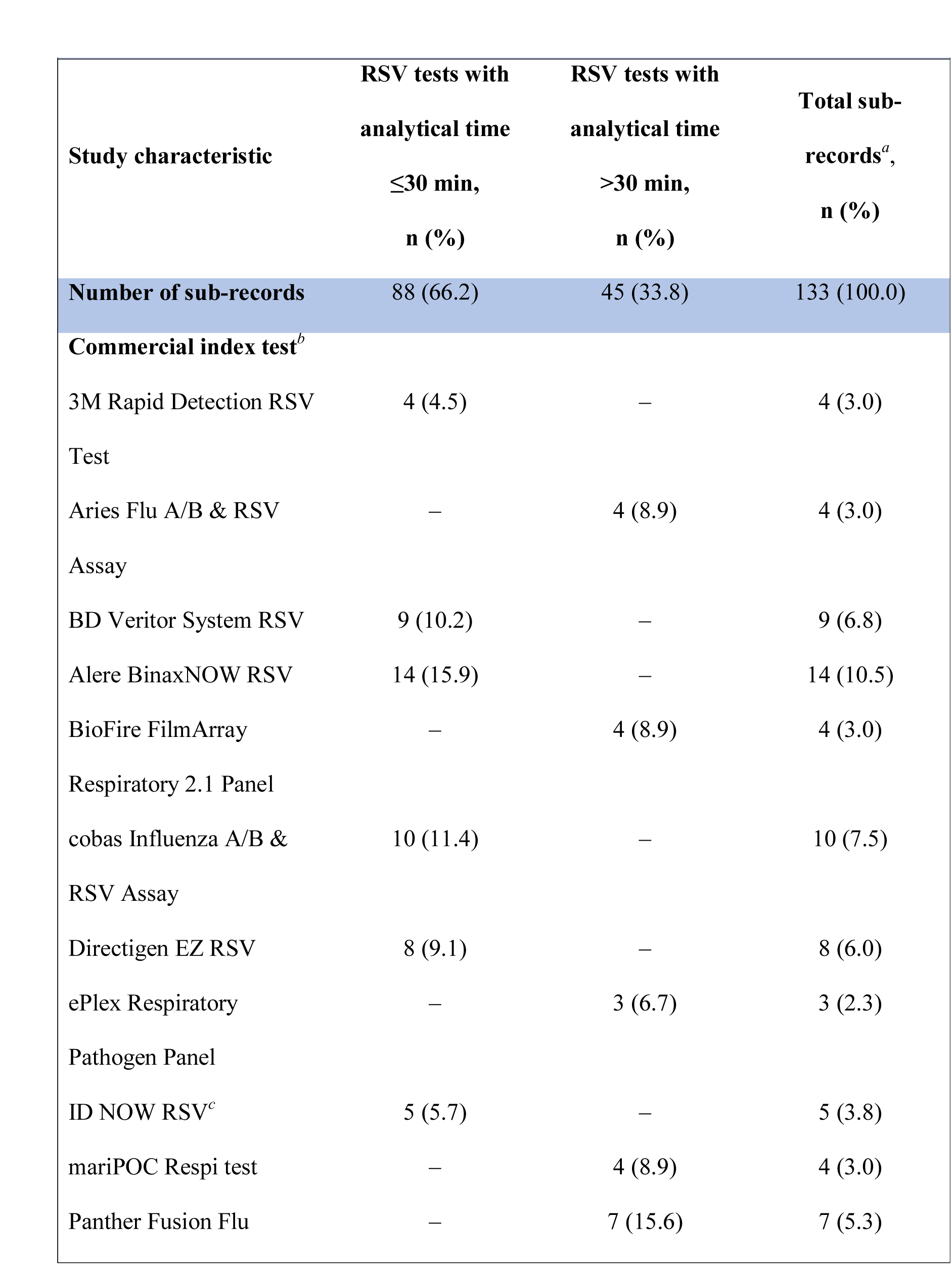
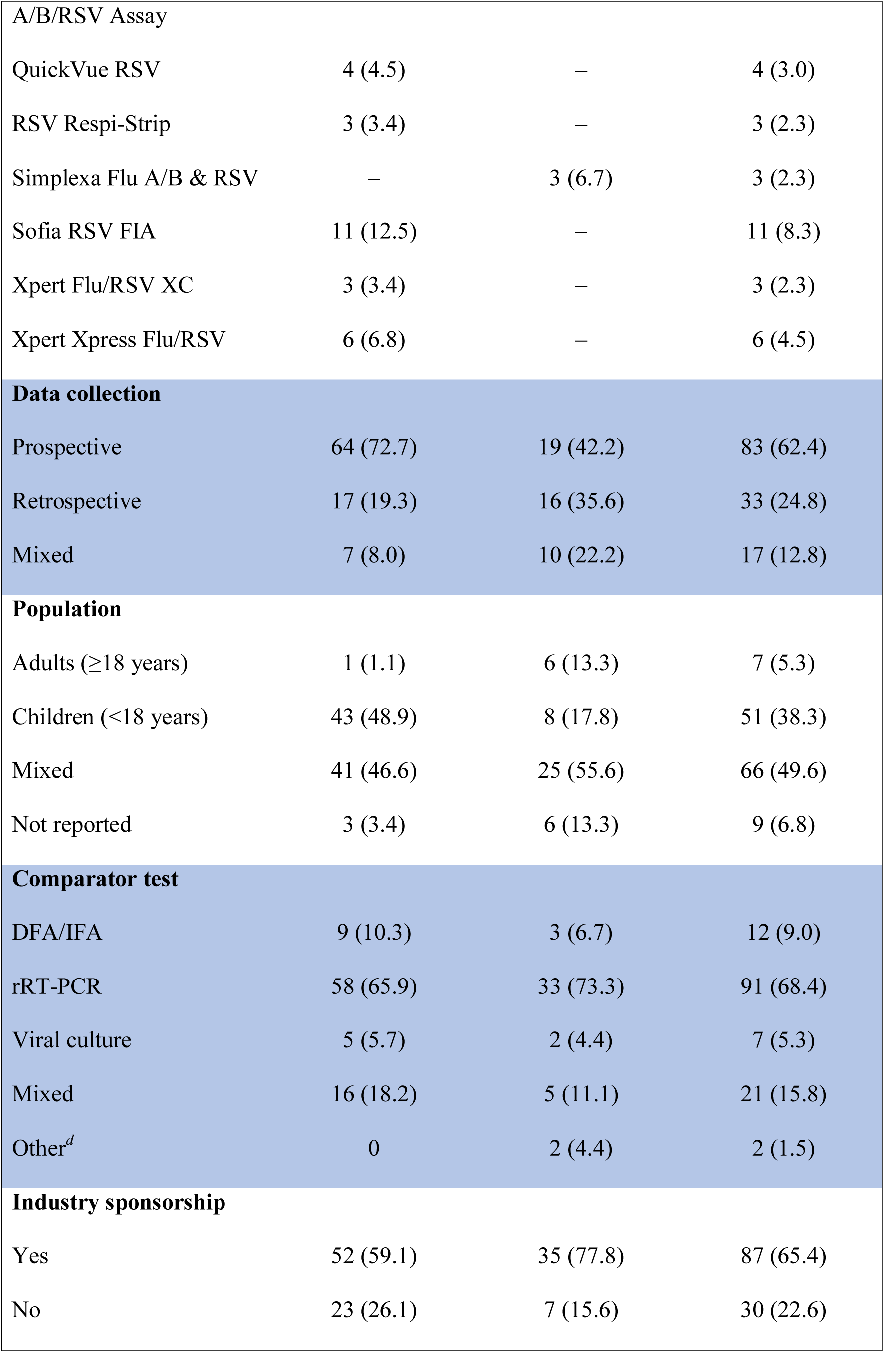
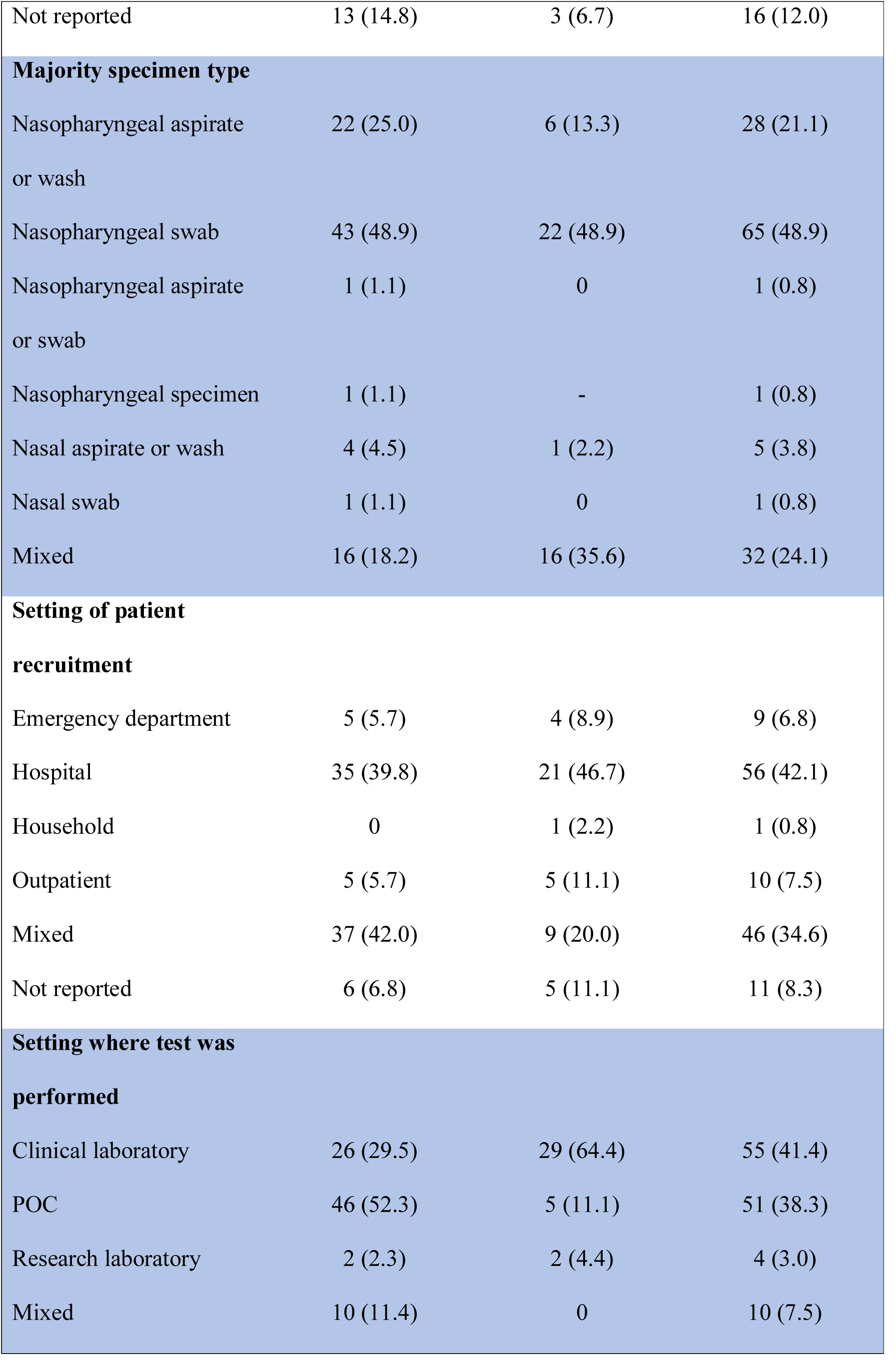
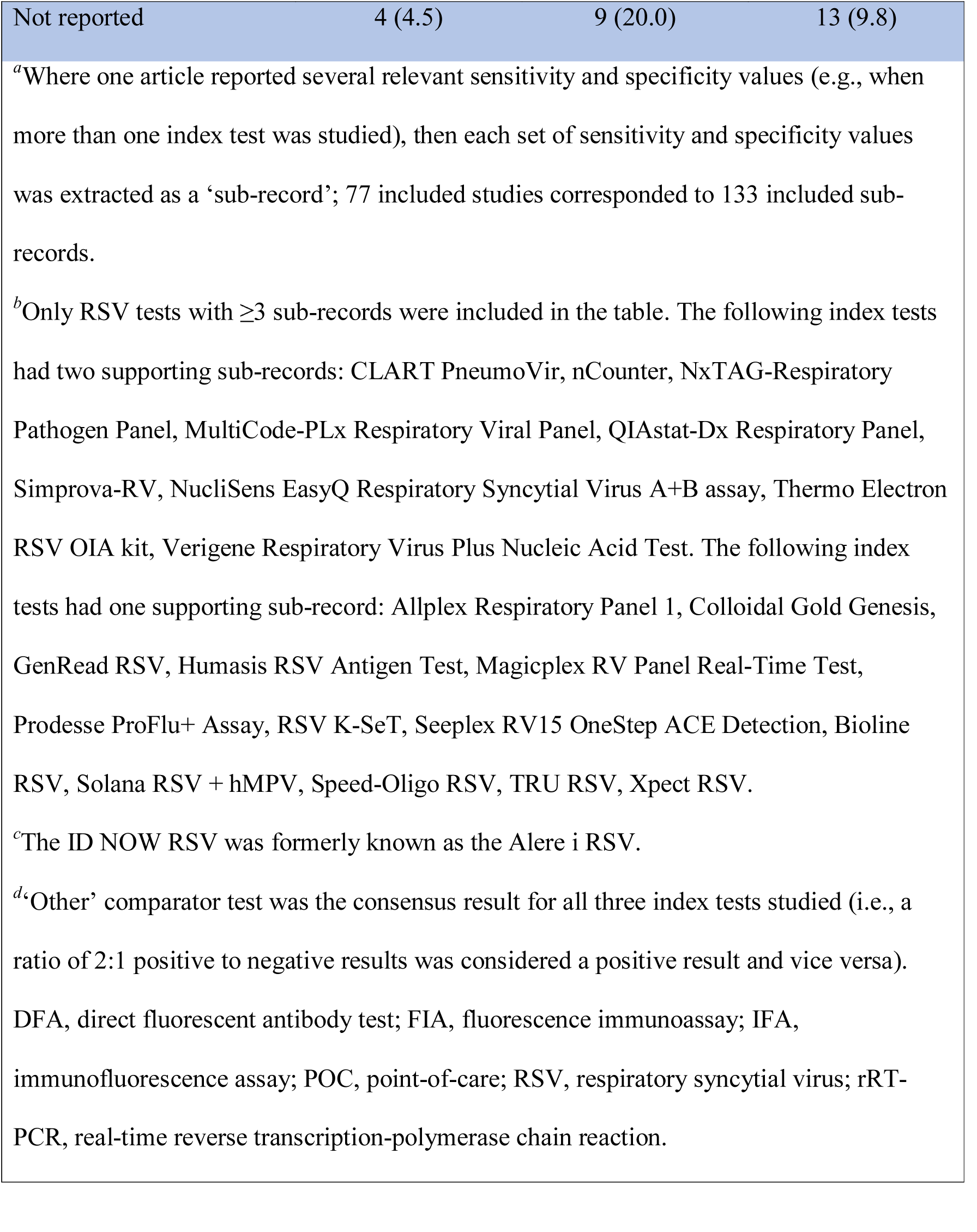
Characteristics of all included studies.

**Fig. 2.**
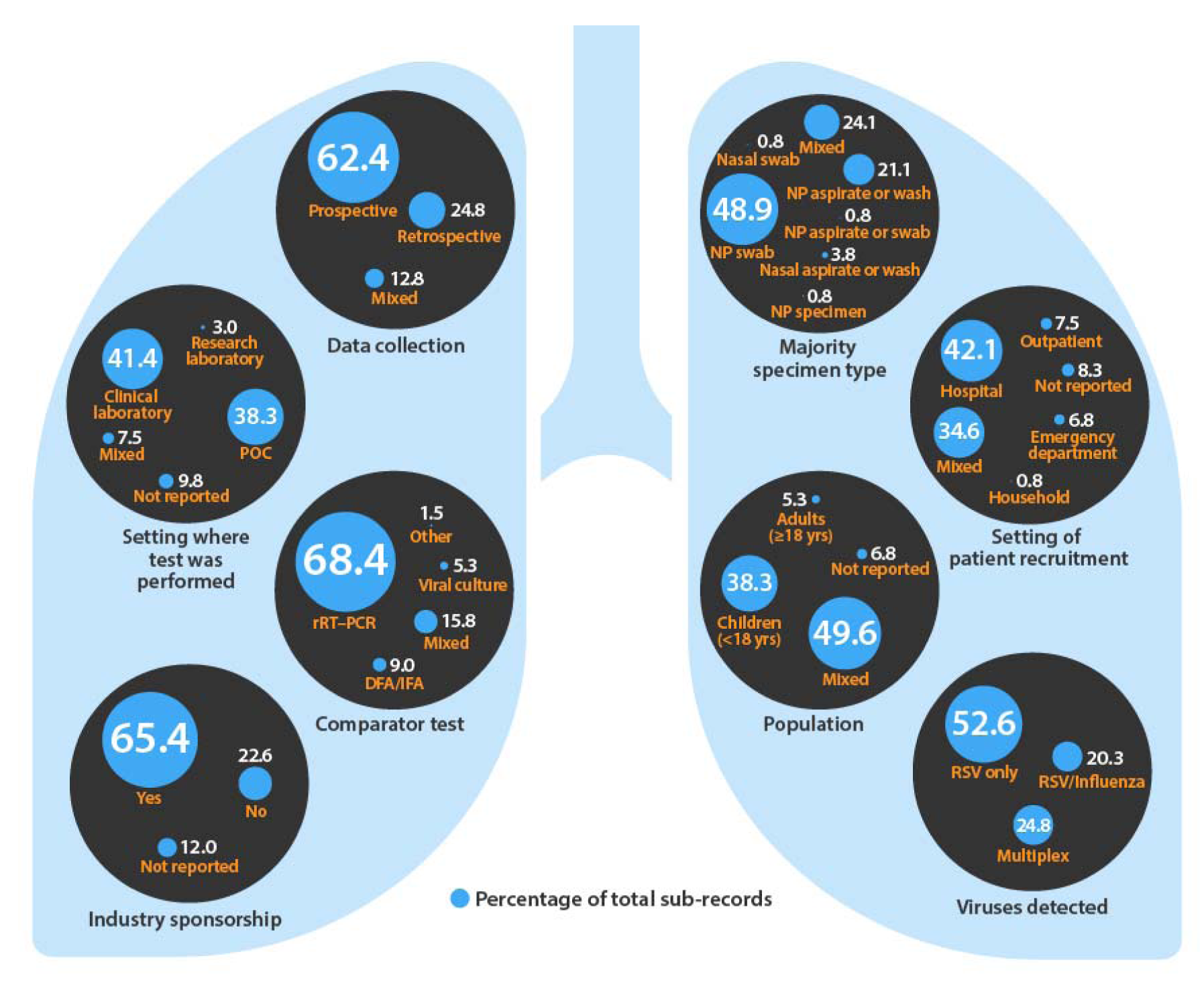
Trends in study design, patient, and setting characteristics in studies included in the review. DFA, direct fluorescent antibody test; IFA, immunofluorescence assay; NP, nasopharyngeal; POC, point-of-care; RSV, respiratory syncytial virus; rRT-PCR, real-time reverse transcription-polymerase chain reaction.

For RSV tests with analytical time ≤30 min, most were performed at the POC (52.3%), whereas RSV tests with analytical time >30 min were predominantly conducted in the clinical laboratory (64.4%) (Table 1). Regarding knowledge gaps in the literature, there was a notably small percentage of studies conducted in adult-only patients (5.3%), few conducted in outpatient settings (7.5%) and only one study was conducted in a household setting (Table 1).

### Sensitivity and Specificity of Antigen- Vs Molecular-Based RSV Tests

RSV tests with analytical time ≤30 min had a greater variability in published sensitivity values (25.7–100%; Table 2) relative to RSV tests with analytical time >30 min (62.5–100%; Table 2); this is partially reflective of the different assays (antigen and molecular) used for RSV tests with analytical time ≤30 min. The range of specificity values was similar for RSV tests with analytical time ≤30 min (80.3–100%) relative to RSV tests with analytical time >30 min (77.0–100%; Table 2).

**Table 2.** Sensitivity and specificity of RSV tests by assay technology in all included.

| Table 2. Sensitivity and specificity of RSV tests by assay technology in all included studies. |  |  |  |
| --- | --- | --- | --- |
| Method | RSV tests with analytical time<br>≤30 min (n = 88) |  | RSV tests with<br>analytical time<br>>30 min (n = 45) |
|  | Antigen | Molecular | Molecular |
| Number of sub-records, n (%) | 61 (69.3) | 27 (30.7) | 41 (91.1)<sup>a</sup> |
| **Overall** |  |  |  |
| Sensitivity, % | 25.7–100 | 66.7–100 | 62.5–100 |
| Specificity, % | 80.3–100 | 94.3–100 | 77.0–100 |
| **Age** |  |  |  |
| <18 years |  |  |  |
| Sensitivity, % | 25.7–97.6 | 84.3–100 | 74.3–100 |
| Specificity, % | 80.3–100 | 94.3–100 | 97.8–100 |
| ≥18 years |  |  |  |
| Sensitivity, %<sup>b</sup> | - | - | 62.5–100 |
| Specificity, %<sup>b</sup> | - | - | 98.9–100 |
| Mixed age |  |  |  |
| Sensitivity, % | 57.5–100 | 77.8–100 | 63.2–100 |
| Specificity, % | 91.8–100 | 94.7–100 | 77.0–100 |
| **Setting** |  |  |  |
| Inpatient |  |  |  |
| Sensitivity, % | 25.7–95.2 | 98.1–100 | 80.4–100 |
| Specificity, % | 80.3–100 | 94.3–99.4 | 91.5–100 |

| Emergency department/outpatient |  |  |  |
| --- | --- | --- | --- |
| Sensitivity, % | 67.8–97.6 | 93–100 | 62.5–100 |
| Specificity, % | 97.6–99.6 | 96–100 | 97.8–100 |
| <sup>a</sup>The mariPOC Respi test (n = 4 sub-records) was included in the RSV tests with analytical time >30 min category as the final result is available after 2 hours. The sensitivity and specificity values were not included in the table as there were insufficient data to produce a range of sensitivity or specificity values for relevant patient and setting characteristics.<br><sup>b</sup>Only one study assessed an RSV test with analytical time ≤30 min in an adult (≥18 years) only population; therefore, it was not possible to produce a range of sensitivity or specificity values.<br>RSV, respiratory syncytial virus. |  |  |  |

Of RSV tests with analytical time ≤30 min, 70.0% (14/20) were antigen-based, and 30.0% (6/20) were molecular tests (Supplemental Table 2). Overall, molecular RSV tests with analytical time ≤30 min had an overall higher sensitivity (66.7–100%) and specificity (94.3–100%), relative to antigen-based RSV tests with analytical time ≤30 min (sensitivity: 25.7–100% and specificity: 80.3–100%; Table 2). This trend for higher diagnostic accuracy in molecular- vs antigen-based RSV tests with analytical time ≤30 min was preserved in all but one of the categories (specificity in the emergency department/outpatient setting) when the sensitivity and specificity ranges were broken down by patient age and setting in which the test was carried out (Table 2).

The sensitivity values of molecular-based tests was highest for those that detected RSV only (93–100%), followed by RSV and influenza (66.7–100%), then multiplex (≥3 viruses detected) platforms (62.5–100%; all Supplemental Table 3). Such summary statistics should be interpreted with caution given the differences in sensitivity between different tests with similar modalities and analytical times e.g., the cobas^®^ Influenza A/B & RSV Assay (Roche Diagnostics International Ltd) and the Xpert Xpress Flu/RSV (Cepheid) are both molecular-based tests that detect RSV and influenza in ≤30 min; however, the sensitivity range reported in the literature for each test is 94.2–100.0% and 66.7–98.1%, respectively (Supplemental Table 4).

### Sensitivity and Specificity of CLIA-Waived Tests for RSV

There was a variety of sensitivity and specificity ranges for the 14 RSV tests included in this review that were assessed under CLIA guidance (Fig. 3). Sensitivity and specificity values for all RSV tests included in the review are shown in Supplemental Table 4. For the CLIA-waived RSV tests, there was a wide range of published sensitivity (25.7–100%) and specificity values (86.8–100%; Table 3). The test with the highest range of sensitivity values was the cobas Influenza A/B & RSV Assay (94.2% [95% confidence interval (CI) 87.9–97.9] – 100.0% [95% CI 96.07–100.0]; Table 3) and the test with the highest range of specificity values was the Xpert Xpress Flu/RSV (98.1% [95% CI 96.6–99.0] – 100% [95% CI 99.7–100]; Table 3).

**Fig. 3.**
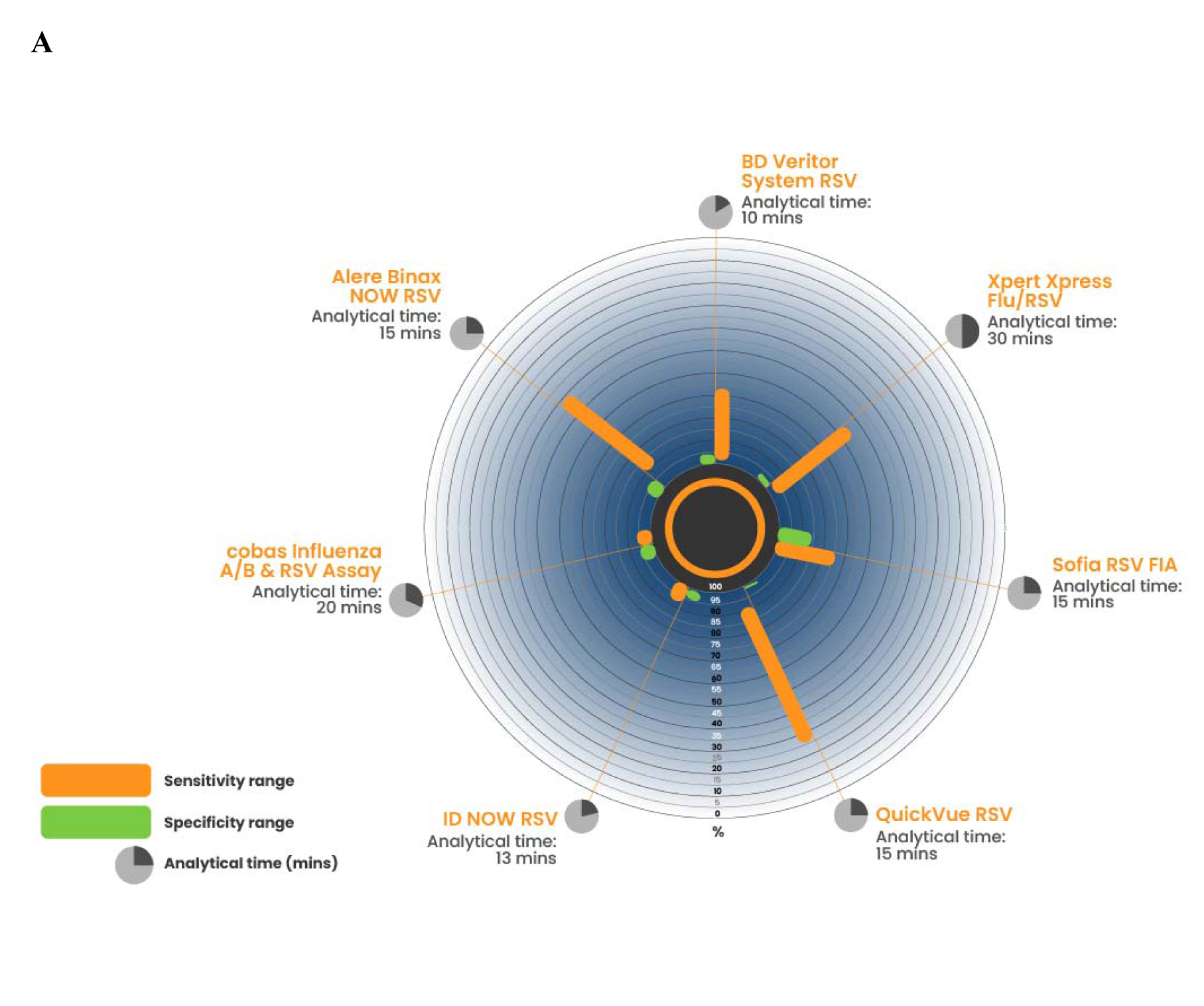
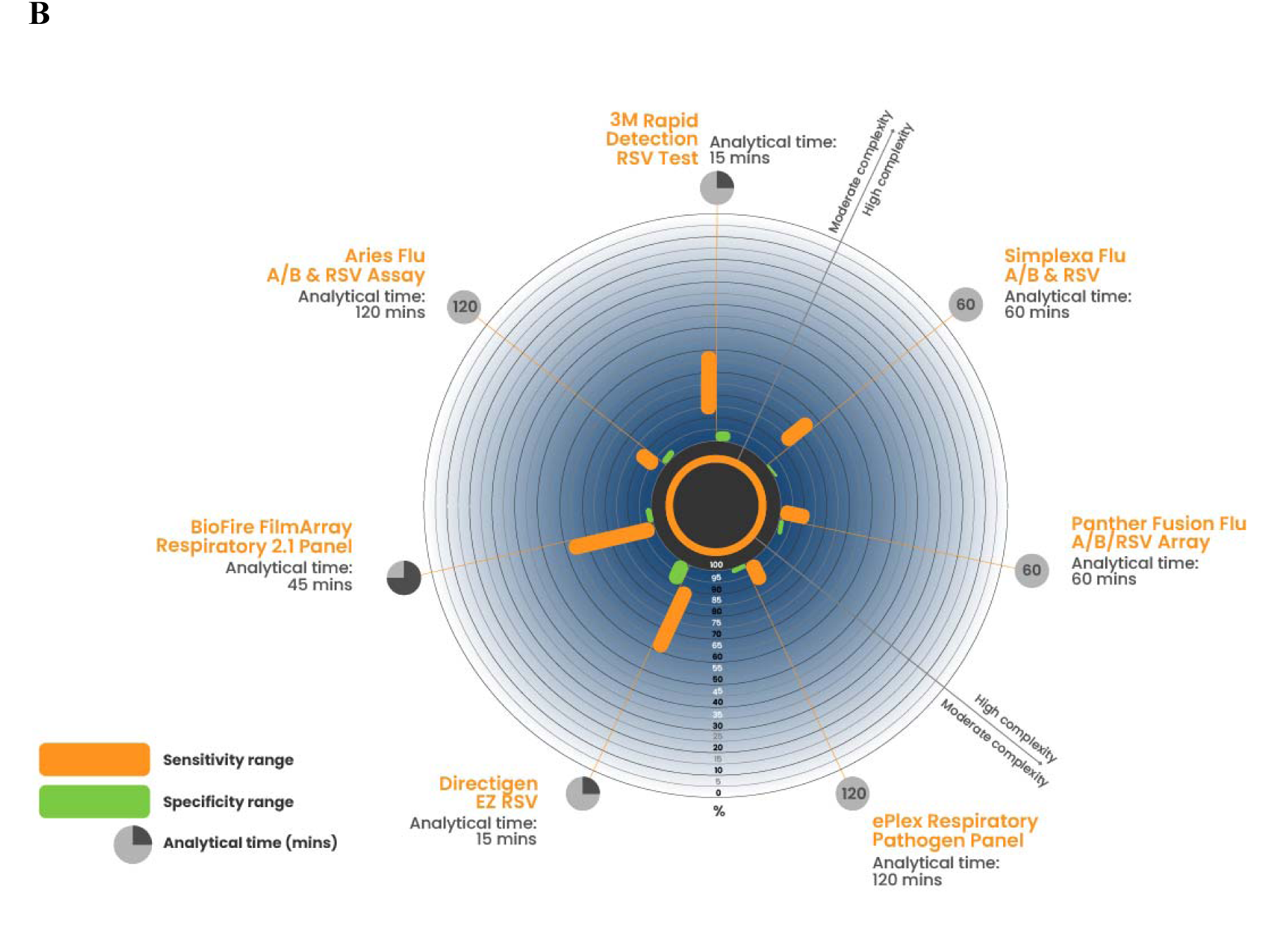
Sensitivity and specificity of RSV tests under CLIA guidance. The sensitivity, specificity and analytical time are shown for the RSV tests included in this review that were CLIA-waived (A) and classed as moderate/high complexity (B). RSV, respiratory syncytial virus.

**Table 3.**
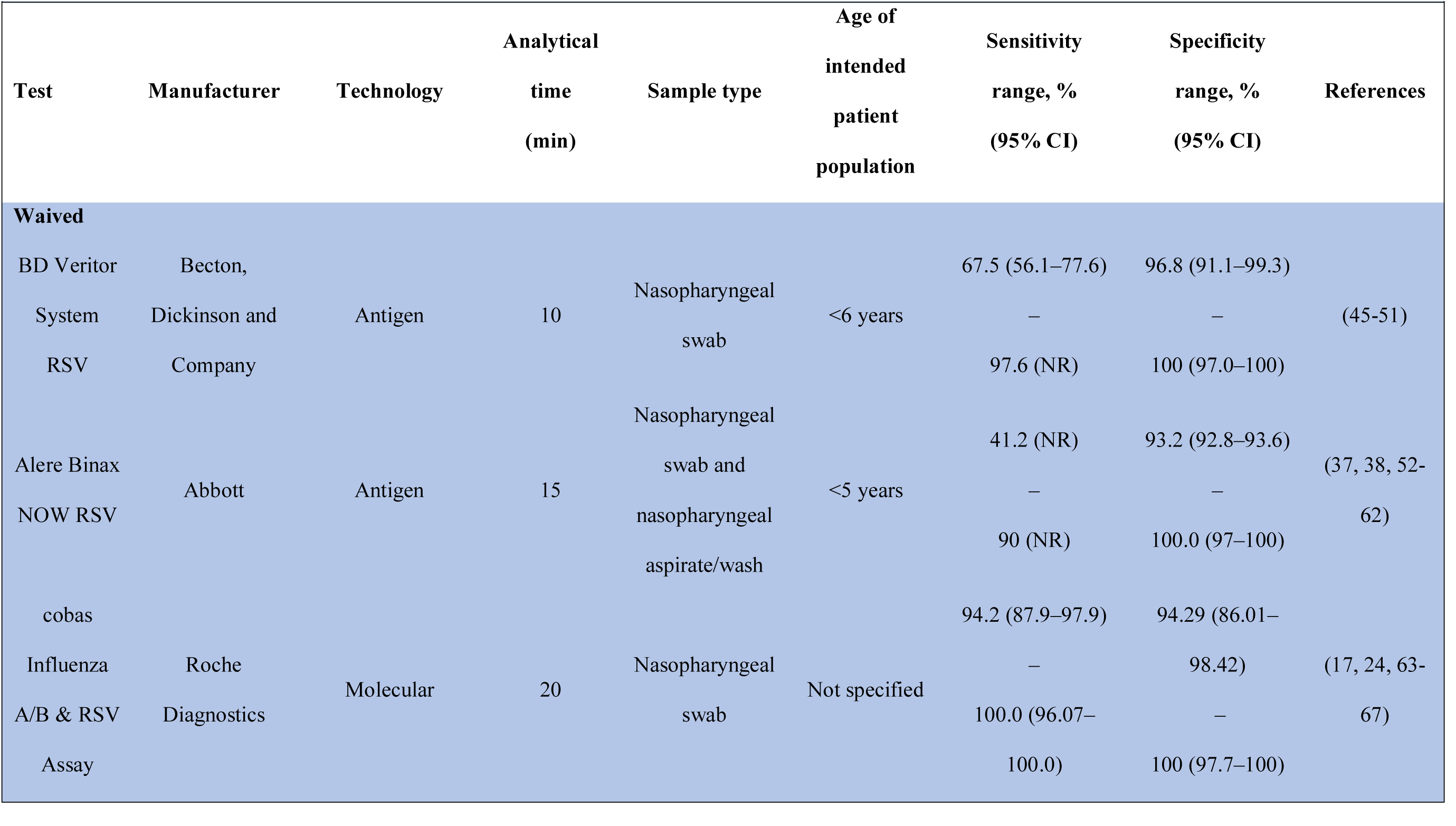
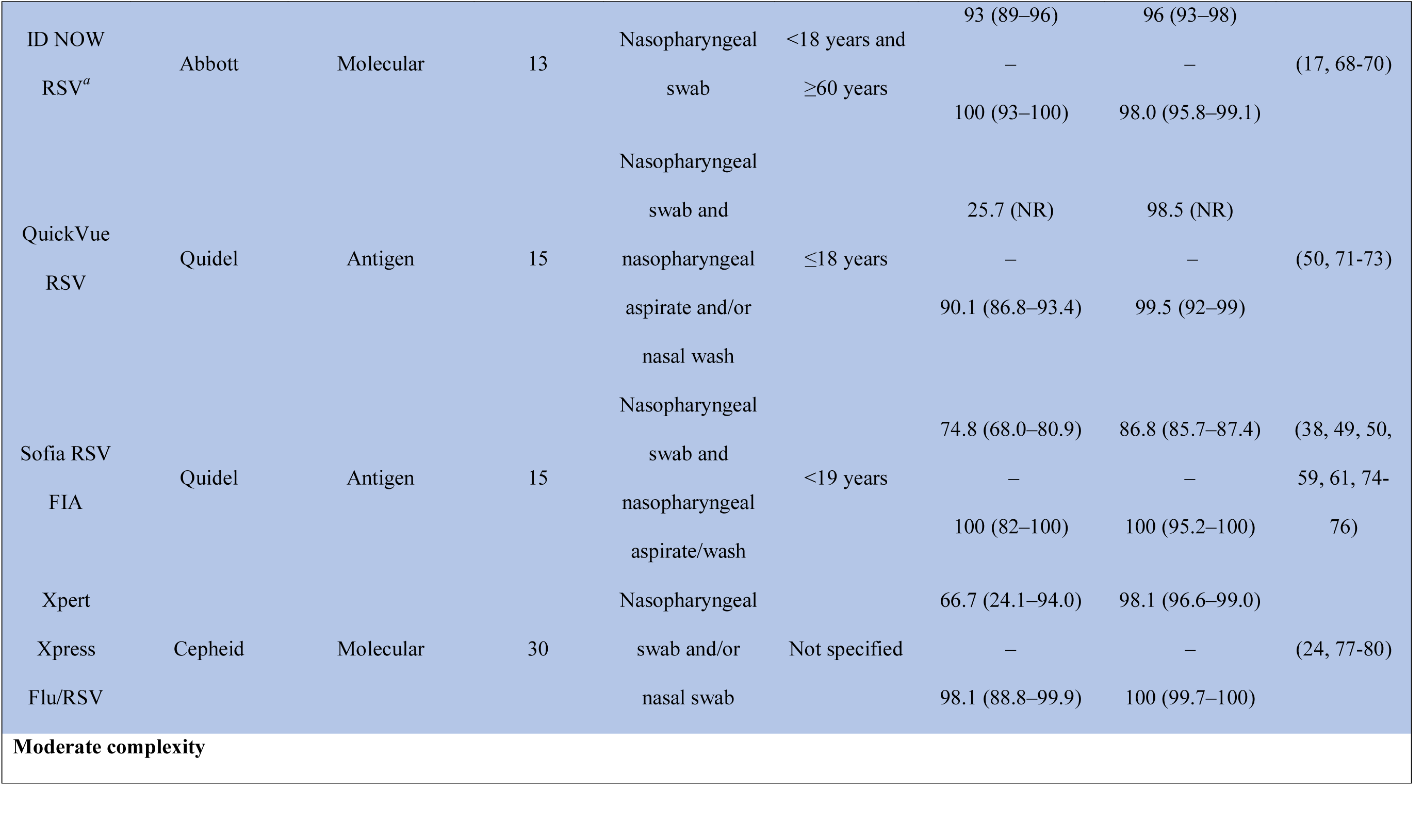
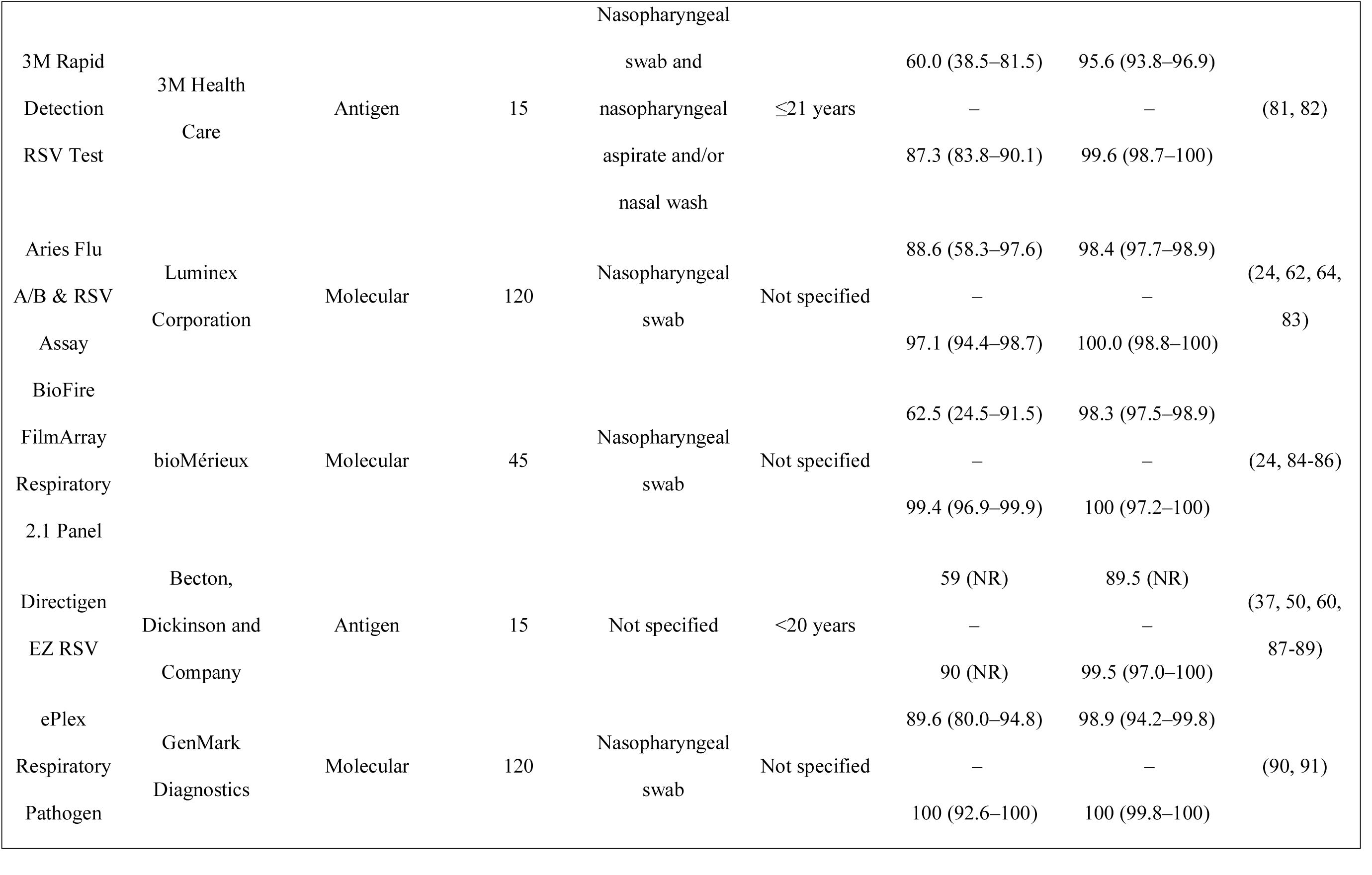
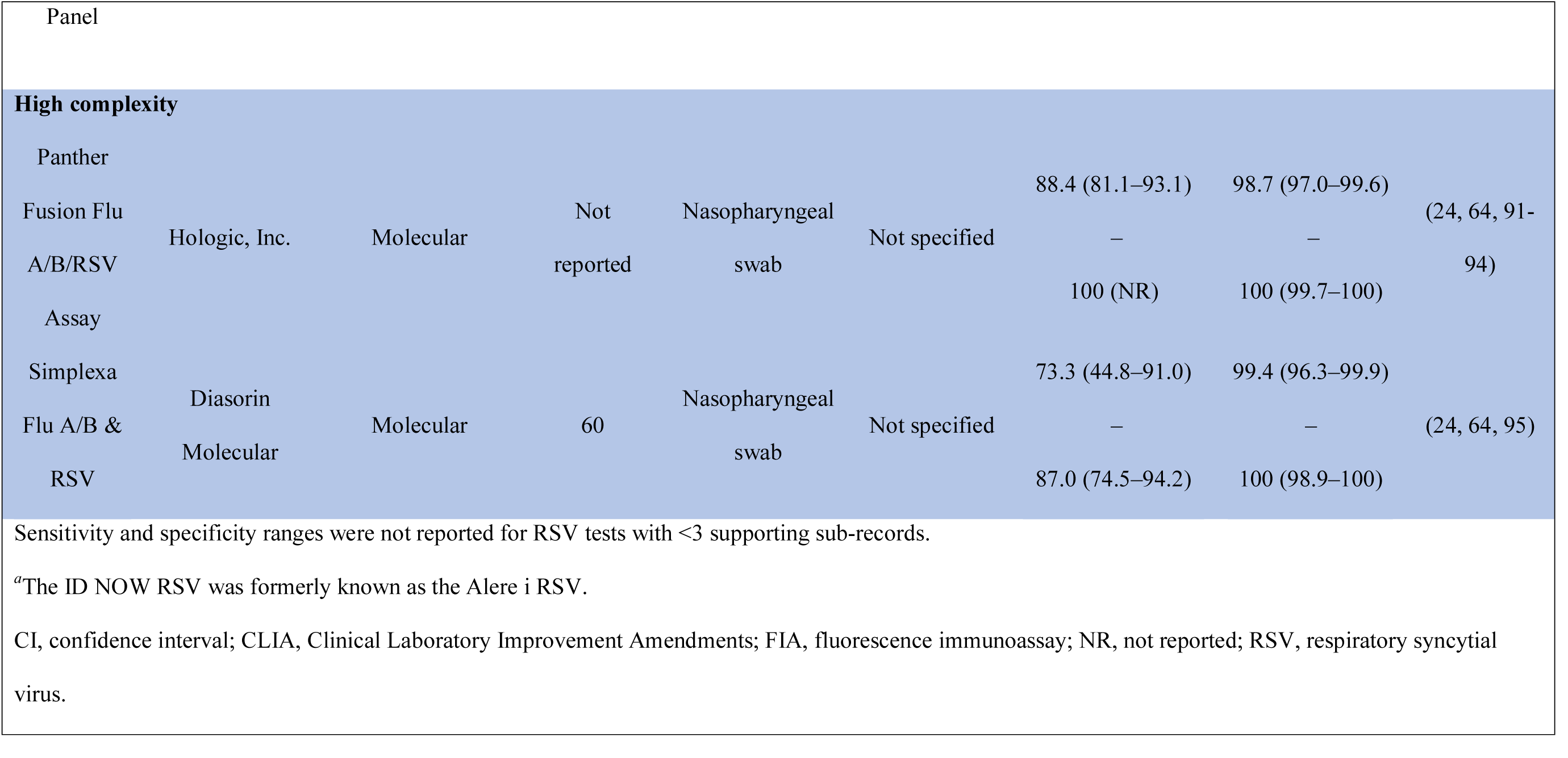
Sensitivity and specificity of RSV tests reviewed under CLIA guidance only.

## DISCUSSION

This scoping literature review summarized sensitivity and specificity values in the peer-reviewed literature for commercially available sample-to-answer tests for RSV. We identified a knowledge gap for studies of RSV tests conducted in adult-only populations or in outpatient or household settings. Overall, RSV tests with analytical time ≤30 min had a greater variability in published sensitivity values relative to RSV tests with analytical time >30 min, which could be partially attributed to the different diagnostic tools (antigen vs molecular) used. Molecular-based rapid RSV tests had higher sensitivity and specificity ranges than antigen-based RSV tests, which aligns with CDC guidance to use molecular testing for RSV where available (11).

The results from this scoping literature review showed a notable gap in studies of diagnostic accuracy of RSV tests in adults. Utilizing viral diagnostic testing in adult patients presenting with symptoms of acute respiratory infection would improve current surveillance efforts and allow for efficient triage and treatment decisions (e.g., local infection control guidance could be followed in a timely manner). This could be particularly important for elderly patients who are at a higher risk of hospitalization and death from RSV infection compared with younger adults (30). Testing strategies for SARS-CoV-2 developed in response to the global COVID- 19 pandemic have undoubtedly brought diagnostics closer to the patient. With respect to RSV, this review identified few published studies available on the diagnostic accuracy of tests in outpatient or household settings. The nasopharyngeal shedding of RSV rapidly decreases 1–3 days after the onset of symptoms (31); therefore, accessible at-home or POC testing could be a valuable tool in timely infection control.

Approximately half of all studies included in this review used a nasopharyngeal swab as the majority specimen type. Notably, it has been shown that the diagnostic accuracy of some types of RSV tests is dependent upon sample type. The sensitivity and specificity of immunofluorescence-based RSV tests is higher in nasopharyngeal aspirates, relative to nasal swabs (32, 33). However, there is no difference in test performance between aspirate and swab specimens when using molecular-based RSV testing (32, 34). Additionally, mid-turbinate nasal swabs have been shown to have a comparative viral load to nasopharyngeal swabs in infants <2 years old (35) and are equally sensitive for the diagnosis of multiple respiratory viruses in adults (36). The advantages of using a nasal swab rather than a nasopharyngeal aspirate/swab are that it is less invasive for the patient and easier for clinical staff to transport (32, 34).

The gold standard for RSV testing, rRT-PCR, was the most used comparator assay across all the studies included in this review. The use of different reference standards has been shown to affect the calculated sensitivity and specificity value of an index test; for example, a significant increase in test sensitivity has been reported in the literature when immunofluorescence is used as the reference standard compared with rRT-PCR (37).

A systematic review and meta-analysis of the sensitivity and specificity of RSV rapid antigen-based tests by Chartrand et al. reported a pooled sensitivity and specificity of 80% (95% CI 76–83) and 97% (95% CI 96–98), respectively (37). In addition, there was a large disparity observed in sensitivity of RSV tests between studies in pediatric patients (81% [95% CI 78–84]) and in adults (29% [95% CI 11–48]). In contrast, a systematic review by Bruning et al. reported that age did not affect diagnostic accuracy of RSV tests; however, this analysis only focused on three rapid RSV tests (BD Veritor System RSV, Sofia RSV FIA, and Alere BinaxNOW RSV) (38). Furthermore, while RSV rapid antigen-based tests are thought to be useful for diagnosis in infants, sensitivity values as low as 7.6% have been reported for a particular brand of rapid antigen-based test in this age group (39).

In some clinical contexts, the use of multiplex tests for more than one respiratory virus may increase efficiency in triaging patients presenting with symptoms of a respiratory tract infection. Young et al. compared the turnaround time for two commercial brands of rapid tests for influenza A and B and RSV. The turnaround time for the ID NOW RSV assay and the ID NOW Influenza A and B assay was 6.4–15.8 min per test result vs 21.3–22.0 min for the combined cobas Influenza A/B & RSV Assay (40). In addition to considering ‘time to result’ for multiplex tests, users should also pay close attention to hands-on time when implementing a new assay. Multiplex RSV tests such as the BioFire FilmArray Respiratory 2.1 Panel and the ePlex Respiratory Pathogen Panel, included in this review, can detect >20 infectious respiratory pathogens; however, these tests are not CLIA-waived and have a longer turnaround time but may be extremely valuable in patients with severe disease where rapid identification of the causative agent(s) in a simultaneous manner may be beneficial. In addition, discrepancies in sensitivity between multiplex and RSV only rRT-PCR tests have been reported in the literature; this could result in varying thresholds for different respiratory viruses between different brands of multiplex rRT-PCR tests (41). The findings from our review also showed that there are differences in the sensitivity and specificity values reported for tests for RSV only relative to multiplex tests.

The evidence outlined in this paper highlights the need for healthcare professionals to consider the spectrum of respiratory disease, not just SARS-CoV-2 or influenza, and consider how viral diagnostic testing could inform their patient management and treatment decisions. If clinicians do not test for RSV, it leads to selection bias and potentially an underestimation of the prevalence of the virus. Most importantly, it could lead to inappropriate treatment for the patient. Healthcare professionals should assess the benefits and drawbacks of each RSV testing method and decide which would be most appropriate in their practice. Factors to consider include the site of testing, the location of the testing instrument, the age and immune status of the individual being tested, end user of the test, where the test results will be analyzed, the clinical significance of the results, implications for infection control, and the added value of a combination test result (40, 42).

One strength of this scoping review is its comprehensive and structured search strategy, which has maximized the capture of relevant information. In addition, this review has considered a broad spectrum of molecular and non-molecular RSV tests with different analytical times. An inherent limitation of scoping reviews is that the data synthesis is based on the values extracted from any given study; therefore, results may not be comparable in terms of methodology, limiting the meaningful conclusions that can be drawn from differences in the sensitivity and specificity between RSV tests from different studies. However, the purpose of this scoping review was not to analyze but to summarize the published data available.

Future research should assess the sensitivity and specificity of RSV tests in adult populations and in outpatient and household settings. In addition, studies should control for selection bias and adjust for differences in settings where the RSV test was performed, seasonality, and staff utilization of RSV tests. The use of POC testing for influenza and RSV across four centers in Denmark resulted in a significant reduction in antibiotic prescription and median hospitalization time in adults (44.3 hours) and children (14.2 hours), there was also an increase in the use of antiviral treatment in adults only (18). These positive results indicate that further studies are warranted to explore the effects of testing for RSV on patient outcomes (43).

In conclusion, different clinical situations (e.g., the clinical laboratory of a large hospital vs an outpatient clinic) will require different diagnostic solutions. Given the higher sensitivity and specificity of molecular-based testing over antigen-based modalities for RSV infection, rRT-PCR tests should be considered for first-line use when possible. By summarizing the sensitivity and specificity data available in the peer-reviewed literature for commercially available RSV tests, this review provides a reference point for healthcare professionals to investigate which test is suitable for their practice. Presently, there are several monoclonal antibodies and vaccines in development for RSV prevention and some promising anti-viral therapeutic agents for RSV treatment (44). The concurrent use of viral diagnostic testing will be become increasingly important to identify the effectiveness and appropriateness of these products in the future.

## Data Availability

The data supporting this review were derived from publicly available databases.

## Acknowledgments

The authors would like to acknowledge Melanie Yarbrough (Washington University School of Medicine, St. Louis, MO, USA) for her valuable input on the design of this literature review. Support for study design and data analysis was provided by Heather Small, PhD, Sophie Lavelle, MSc, and Claire Snowball, MSc, of Ashfield MedComms, Macclesfield, UK, an Ashfield Health Company, and was funded by Roche Diagnostics International Ltd, Rotkreuz, Switzerland. Medical writing support for the development of this manuscript, under the direction of the authors, was provided by Heather Small, PhD, of Ashfield MedComms, Macclesfield, UK, an Ashfield Health Company, and was funded by Roche Diagnostics International Ltd, Rotkreuz, Switzerland. COBAS is a trademark of Roche. All other product names and trademarks are the property of their respective owners.

## Funding

This review was funded by Roche Diagnostics International Ltd, Rotkreuz, Switzerland.

## Ethics approval

Ethical approval was not applicable for this review.

## Author contributions

All authors contributed to the conceptualization of the study, the writing of the original draft and reviewing and editing of subsequent manuscript drafts. DIB contributed to the formal analysis and methodology of the study. JPD contributed to the data curation and supervision of the study.

## Conflicts of interest

DIB is a paid consultant for Roche Diagnostics. AM has received research grants from NIH, Janssen and Merck and fees for participation in advisory boards from Janssen, Sanofi-Pasteur, Merck and Roche Diagnostics. BR has received institutional funding support for research and fees for participation in advisory boards from Roche Diagnostics and Quidel. CWW has received grants from DARPA, NIH/ARLG, NIH/VTEU and Sanofi, consultant fees from bioMerieux, Biofire, Giner, Biomeme, FHI Clinical, Arena Pharmaceuticals, SeLux Diagnostics and Karius, fees from participation in a data safety monitoring board from Janssen, and fees from participation in advisory boards from Regeneron and IDbyDNA; has the following patents: biomarkers for the molecular classification of bacterial infection (issued), methods to diagnose and treat acute respiratory infections (pending), gene expression signatures useful to predict or diagnose sepsis (pending), host based molecular signatures of human infection with SARs-CoV-2 (COVID-19) (pending) and methods of identifying infectious disease and assays for identifying infectious disease (issued); holds stocks in Predigen Inc (equity, founder); is employed by Duke University and Durham VA Health Care System; and is the principal investigator for RADx UP testing Core. JPD is an employee of Roche Diagnostics Corporation.

## Data availability

The data supporting this review were derived from publicly available databases.

## References

1. Paramore LC, Ciuryla V, Ciesla G, Liu L. Economic impact of respiratory syncytial virus-related illness in the US: an analysis of national databases. Pharmacoeconomics 2004;22(5):275–84.

2. Li X, Willem L, Antillon M, Bilcke J, Jit M, Beutels P. Health and economic burden of respiratory syncytial virus (RSV) disease and the cost-effectiveness of potential interventions against RSV among children under 5 years in 72 Gavi-eligible countries. BMC Med 2020;18(1):82.

3. Korsten K, Adriaenssens N, Coenen S, Butler C, Ravanfar B, Rutter H, et al. Burden of respiratory syncytial virus infection in community-dwelling older adults in Europe (RESCEU): an international prospective cohort study. Eur Respir J 2020:57(4):2002688.

4. Malosh RE, Martin ET, Callear AP, Petrie JG, Lauring AS, Lamerato L, et al. Respiratory syncytial virus hospitalization in middle-aged and older adults. J Clin Virol 2017;96:37–43.

5. Falsey AR, Hennessey PA, Formica MA, Cox C, Walsh EE. Respiratory syncytial virus infection in elderly and high-risk adults. N Engl J Med 2005;352(17):1749–59.

6. Ackerson B, An J, Sy LS, Solano Z, Slezak J, Tseng HF. Cost of hospitalization associated with respiratory syncytial virus infection versus influenza infection in hospitalized older adults. J Infect Dis 2020;222(6):962–6.

7. Borchers AT, Chang C, Gershwin ME, Gershwin LJ. Respiratory syncytial virus--a comprehensive review. Clin Rev Allergy Immunol 2013;45(3):331–79.

8. Falsey AR. Respiratory syncytial virus infection in adults. Semin Respir Crit Care Med 2007;28(2):171–81.

9. Midgley CM, Haynes AK, Baumgardner JL, Chommanard C, Demas SW, Prill MM, et al. Determining the seasonality of respiratory syncytial virus in the United States: the impact of increased molecular testing. J Infect Dis 2017;216(3):345–55.

10. van de Pol AC, Wolfs TFW, Jansen NJG, van Loon AM, Rossen JWA. Diagnostic value of real-time polymerase chain reaction to detect viruses in young children admitted to the paediatric intensive care unit with lower respiratory tract infection. Critical Care 2006;10(2):R61.

11. Centers for Disease Control and Prevention. Respiratory syncitial virus for healthcare providers. https://www.cdc.gov/rsv/clinical/index.html (Accessed October 01, 2021).

12. Lee N, Walsh EE, Sander I, Stolper R, Zakar J, Wyffels V, et al. Delayed diagnosis of respiratory syncytial virus infections in hospitalized adults: individual patient data, record review analysis and physician survey in the United States. J Infect Dis 2019;220(6):969–79.

13. Bont L, Checchia PA, Fauroux B, Figueras-Aloy J, Manzoni P, Paes B, et al. Defining the epidemiology and burden of severe respiratory syncytial virus infection among infants and children in Western countries. Infect Dis Ther 2016;5(3):271–98.

14. Walsh EE. Respiratory syncytial virus infection: an illness for all ages. Clin Chest Med 2017;38(1):29–36.

15. Staadegaard L, Caini S, Wangchuk S, Thapa B, de Almeida WAF, de Carvalho FC, et al. The global epidemiology of RSV in community and hospitalized care: findings from 15 countries. Open Forum Infect Dis 2021;8(7):ofab159.

16. Alchikh M, Conrad T, Hoppe C, Ma X, Broberg E, Penttinen P, et al. Are we missing respiratory viral infections in infants and children? Comparison of a hospital-based quality management system with standard of care. Clin Microbiol Infect 2019;25(3):380.e9-.e16.

17. Leonardi GP. Evaluation of rapid, molecular-based assays for the detection of respiratory syncytial virus. Intervirology 2019;62(3-4):112–5.

18. Schneider UV, Holm MKA, Bang D, Petersen RF, Mortensen S, Trebbien R, et al. Point-of-care tests for influenza A and B viruses and RSV in emergency departments - indications, impact on patient management and possible gains by syndromic respiratory testing, Capital Region, Denmark, 2018. Euro Surveill 2020;25(44):1900430.

19. Baker RE, Park SW, Yang W, Vecchi GA, Metcalf CJE, Grenfell BT. The impact of COVID-19 nonpharmaceutical interventions on the future dynamics of endemic infections. Proc Natl Acad Sci U S A 2020;117(48):30547–53.

20. Centers for Disease Control and Prevention. Increased interseasonal respiratory syncytial virus (RSV) activity in parts of the Southern United States. https://emergency.cdc.gov/han/2021/pdf/CDC-HAN-443-Increased-Interseasonal-RSV-Activity-06.10.21.pdf (Accessed June 16, 2021).

21. Agha R, Avner JR. Delayed seasonal RSV surge observed during the COVID-19 pandemic. Pediatrics 2021;148(3):e2021052089.

22. von Hammerstein AL, Aebi C, Barbey F, Berger C, Buettcher M, Casaulta C, et al. Interseasonal RSV infections in Switzerland - rapid establishment of a clinician-led national reporting system (RSV EpiCH). Swiss Med Wkly 2021;151:w30057.

23. Foley DA, Yeoh DK, Minney-Smith CA, Martin AC, Mace AO, Sikazwe CT, et al. The interseasonal resurgence of respiratory syncytial virus in Australian children following the reduction of coronavirus disease 2019-related public health measures. Clin Infect Dis 2021;73(9):e2829–2830.

24. Banerjee D, Kanwar N, Hassan F, Essmyer C, Selvarangan R. Comparison of six sample-to-answer influenza A/B and respiratory syncytial virus nucleic acid amplification assays using respiratory specimens from children. J Clin Microbiol 2018;56(11):e00930–18.

25. Barr R, Green CA, Sande CJ, Drysdale SB. Respiratory syncytial virus: diagnosis, prevention and management. Ther Adv Infect Dis 2019;6:2049936119865798.

26. Centers for Disease Control and Prevention. About CLIA. https://www.cdc.gov/clia/about.html (Accessed October 01, 2021).

27. Centers for Disease Control and Prevention. Test Complexities. https://www.cdc.gov/clia/test-complexities.html (Accessed October 01, 2021).

28. Arksey H, O’Malley L. Scoping studies: towards a methodological framework. Int J Soc Res Methodol 2005;8(1):19–32.

29. Tricco AC, Lillie E, Zarin W, O’Brien KK, Colquhoun H, Levac D, et al. PRISMA Extension for Scoping Reviews (PRISMA-ScR): checklist and explanation. Ann Intern Med 2018;169(7):467–73.

30. Watson A, Wilkinson TMA. Respiratory viral infections in the elderly. Ther Adv Respir Dis 2021;15:1753466621995050.

31. Abels S, Nadal D, Stroehle A, Bossart W. Reliable detection of respiratory syncytial virus infection in children for adequate hospital infection control management. J Clin Microbiol 2001;39(9):3135–9.

32. Sung RYT, Chan PKS, Choi KC, Yeung ACM, Li AM, Tang JW, et al. Comparative study of nasopharyngeal aspirate and nasal swab specimens for diagnosis of acute viral respiratory infection. J Clin Microbiol 2008;46(9):3073–6.

33. Macfarlane P, Denham J, Assous J, Hughes C. RSV testing in bronchiolitis: which nasal sampling method is best? Arch Dis Child 2005;90(6):634.

34. Abu-Diab A, Azzeh M, Ghneim R, Ghneim R, Zoughbi M, Turkuman S, et al. Comparison between pernasal flocked swabs and nasopharyngeal aspirates for detection of common respiratory viruses in samples from children. J Clin Microbiol 2008;46(7):2414–7.

35. Blaschke AJ, McKevitt M, Ampofo K, Lewis T, Chai H, Guo Y, et al. A mid- turbinate swab appears comparable to nasopharyngeal swabs for quantitative detection of RSV in infants. Open Forum Infect Dis 2017;4(Suppl 1):S354–S5.

36. Larios OE, Coleman BL, Drews SJ, Mazzulli T, Borgundvaag B, Green K, et al. Self- collected mid-turbinate swabs for the detection of respiratory viruses in adults with acute respiratory illnesses. PLoS One 2011;6(6):e21335.

37. Chartrand C, Tremblay N, Renaud C, Papenburg J. Diagnostic accuracy of rapid antigen detection tests for respiratory syncytial virus infection: systematic review and meta-analysis. J Clin Microbiol 2015;53(12):3738–49.

38. Bruning AHL, Leeflang MMG, Vos J, Spijker R, de Jong MD, Wolthers KC, et al. Rapid tests for influenza, respiratory syncytial virus, and other respiratory viruses: a systematic review and meta-analysis. Clin Infect Dis 2017;65(6):1026–32.

39. Zuurbier RP, Bont LJ, Langedijk AC, Hamer M, Korsten K, Drysdale SB, et al. Low Sensitivity of BinaxNOW RSV in Infants. J Infect Dis 2020;222(Suppl 7).

40. Young S, Phillips J, Griego-Fullbright C, Wagner A, Jim P, Chaudhuri S, et al. Molecular point-of-care testing for influenza A/B and respiratory syncytial virus: comparison of workflow parameters for the ID Now and cobas Liat systems. J Clin Pathol 2020;73(6):328–34.

41. Alchikh M, Conrad T, Hoppe C, Ma X, Broberg E, Penttinen P, et al. Are we missing respiratory viral infections in infants and children? Comparison of a hospital-based quality management system with standard of care. Clin Microbiol Infect 2019;25(3):e9-.e16.

42. Kosack CS, Page A-L, Klatser PR. A guide to aid the selection of diagnostic tests. Bull World Health Organ 2017;95(9):639–45.

43. Bordley WC, Viswanathan M, King VJ, Sutton SF, Jackman AM, Sterling L, et al. Diagnosis and testing in bronchiolitis: a systematic review. Arch Pediatr Adolesc Med 2004;158(2):119–26.

44. Domachowske JB, Anderson EJ, Goldstein M. The future of respiratory syncytial virus disease prevention and treatment. Infect Dis Ther 2021;10(Suppl 1):47–60.

45. Bell JJ, Anderson EJ, Greene WH, Romero JR, Merchant M, Selvarangan R. Multicenter clinical performance evaluation of BD Veritor™ system for rapid detection of respiratory syncytial virus. J Clin Virol 2014;61(1):113–7.

46. Bruning AHL, de Kruijf WB, van Weert H, Willems WLM, de Jong MD, Pajkrt D, et al. Diagnostic performance and clinical feasibility of a point-of-care test for respiratory viral infections in primary health care. Fam Pract 2017;34(5):558–63.

47. Cantais A, Mory O, Plat A, Giraud A, Pozzetto B, Pillet S. Analytical performances of the BD Veritor™ System for the detection of respiratory syncytial virus and influenzaviruses A and B when used at bedside in the pediatric emergency department. J Virol Methods 2019;270:66–9.

48. Jonckheere S, Verfaillie C, Boel A, Van Vaerenbergh K, Vanlaere E, Vankeerberghen A, et al. Multicenter evaluation of BD Veritor System and RSV K-SeT for rapid detection of respiratory syncytial virus in a diagnostic laboratory setting. Diagn Microbiol Infect Dis 2015;83(1):37–40.

49. Kanwar N, Hassan F, Nguyen A, Selvarangan R. Head-to-head comparison of the diagnostic accuracies of BD Veritor™ System RSV and Quidel® Sofia® RSV FIA systems for respiratory syncytial virus (RSV) diagnosis. J Clin Virol 2015;65:83–6.

50. Leonardi GP, Wilson AM, Dauz M, Zuretti AR. Evaluation of respiratory syncytial virus (RSV) direct antigen detection assays for use in point-of-care testing. J Virol Methods 2015;213:131–4.

51. Schwartz RH, Selvarangan R, Zissman EN. BD Veritor System respiratory syncytial virus rapid antigen detection test: point-of-care results in primary care pediatric offices compared with reverse transcriptase polymerase chain reaction and viral culture methods. Pediatr Emerg Care 2015;31(12):830–4.

52. Borek AP, Clemens SH, Gaskins VK, Aird DZ, Valsamakis A. Respiratory syncytial virus detection by Remel Xpect, Binax Now RSV, direct immunofluorescent staining, and tissue culture. J Clin Microbiol 2006;44(3):1105–7.

53. Cruz AT, Cazacu AC, Greer JM, Demmler GJ. Performance of a rapid assay (Binax NOW) for detection of respiratory syncytial virus at a children’s hospital over a 3-year period. J Clin Microbiol 2007;45(6):1993–5.

54. Jung BK, Choi SH, Lee JH, Lee J, Lim CS. Performance evaluation of four rapid antigen tests for the detection of respiratory syncytial virus. J Med Virol 2016;88(10):1720–4.

55. Khanom AB, Velvin C, Hawrami K, Schutten M, Patel M, Holmes MV, et al. Performance of a nurse-led paediatric point of care service for respiratory syncytial virus testing in secondary care. J Infect 2011;62(1):52–8.

56. Liao RS, Tomalty LL, Majury A, Zoutman DE. Comparison of viral isolation and multiplex real-time reverse transcription-PCR for confirmation of respiratory syncytial virus and influenza virus detection by antigen immunoassays. J Clin Microbiol 2009;47(3):527–32.

57. Miernyk K, Bulkow L, DeByle C, Chikoyak L, Hummel KB, Hennessy T, et al. Performance of a rapid antigen test (Binax NOW® RSV) for diagnosis of respiratory syncytial virus compared with real-time polymerase chain reaction in a pediatric population. J Clin Virol 2011;50(3):240–3.

58. Moesker FM, van Kampen JJA, Aron G, Schutten M, van de Vijver D, Koopmans MPG, et al. Diagnostic performance of influenza viruses and RSV rapid antigen detection tests in children in tertiary care. J Clin Virol 2016;79:12–7.

59. Rack-Hoch AL, Laniado G, Hübner J. Comparison of influenza and RSV diagnostic from nasopharyngeal swabs by rapid fluorescent immunoassay (Sofia system) and rapid bedside testing (BinaxNOW) vs. conventional fluorescent immunoassay in a German university children’s hospital. Infection 2017;45(4):529–32.

60. Selvarangan R, Abel D, Hamilton M. Comparison of BD Directigen EZ RSV and Binax NOW RSV tests for rapid detection of respiratory syncytial virus from nasopharyngeal aspirates in a pediatric population. Diagn Microbiol Infect Dis 2008;62(2):157–61.

61. Sun Y, Deng J, Qian Y, Zhu R, Wang F, Tian R, et al. Laboratory evaluation of rapid antigen detection tests for more-sensitive detection of respiratory syncytial virus antigen. Jpn J Infect Dis 2019;72(6):394–8.

62. Voermans JJ, Seven-Deniz S, Fraaij PL, van der Eijk AA, Koopmans MP, Pas SD. Performance evaluation of a rapid molecular diagnostic, MultiCode based, sample-to- answer assay for the simultaneous detection of Influenza A, B and respiratory syncytial viruses. J Clin Virol 2016;85:65–70.

63. Allen AJ, Gonzalez-Ciscar A, Lendrem C, Suklan J, Allen K, Bell A, et al. Diagnostic and economic evaluation of a point-of-care test for respiratory syncytial virus. ERJ Open Res 2020;6(3):00018–2020.

64. Banerjee D, Kanwar N, Hassan F, Lankachandra K, Selvarangan R. Comparative analysis of four sample-to-answer influenza A/B and RSV nucleic acid amplification assays using adult respiratory specimens. J Clin Virol 2019;118:9–13.

65. Gibson J, Schechter-Perkins EM, Mitchell P, Mace S, Tian Y, Williams K, et al. Multi-center evaluation of the cobas(®) Liat(®) Influenza A/B & RSV assay for rapid point of care diagnosis. J Clin Virol 2017;95:5–9.

66. Gosert R, Naegele K, Hirsch HH. Comparing the Cobas Liat Influenza A/B and respiratory syncytial virus assay with multiplex nucleic acid testing. J Med Virol 2019;91(4):582–7.

67. Verbakel JY, Matheeussen V, Loens K, Kuijstermans M, Goossens H, Ieven M, et al. Performance and ease of use of a molecular point-of-care test for influenza A/B and RSV in patients presenting to primary care. Eur J Clin Microbiol Infect Dis 2020;39(8):1453–60.

68. Schnee SV, Pfeil J, Ihling CM, Tabatabai J, Schnitzler P. Performance of the Alere i RSV assay for point-of-care detection of respiratory syncytial virus in children. BMC Infect Dis 2017;17(1):767.

69. Hassan F, Hays LM, Bonner A, Bradford BJ, Franklin R, Jr., Hendry P, et al. Multicenter clinical evaluation of the Alere i respiratory syncytial virus isothermal nucleic acid amplification assay. J Clin Microbiol 2018;56(3):e01777–17.

70. Peters RM, Schnee SV, Tabatabai J, Schnitzler P, Pfeil J. Evaluation of Alere i RSV for rapid detection of respiratory syncytial virus in children hospitalized with acute respiratory tract infection. J Clin Microbiol 2017;55(4):1032–6.

71. Freeman MC, Haddadin Z, Lawrence L, Piya B, Krishnaswami S, Faouri S, et al. Utility of RSV rapid diagnostic assays in hospitalized children in Amman, Jordan. J Med Virol 2020;93:3420–7.

72. Mesquita FDS, Oliveira DBL, Crema D, Pinez CMN, Colmanetti TC, Thomazelli LM, et al. Rapid antigen detection test for respiratory syncytial virus diagnosis as a diagnostic tool. J Pediatr (Rio J) 2017;93(3):246–52.

73. Rath B, Tief F, Obermeier P, Tuerk E, Karsch K, Muehlhans S, et al. Early detection of influenza A and B infection in infants and children using conventional and fluorescence-based rapid testing. J Clin Virol 2012;55(4):329–33.

74. Gomez S, Prieto C, Folgueira L. A prospective study to assess the diagnostic performance of the Sofia(®) Immunoassay for influenza and RSV detection. J Clin Virol 2016;77:1–4.

75. Tran LC, Tournus C, Dina J, Morello R, Brouard J, Vabret A. SOFIA(®)RSV: prospective laboratory evaluation and implementation of a rapid diagnostic test in a pediatric emergency ward. BMC Infect Dis 2017;17(1):452.

76. Tuttle R, Weick A, Schwarz WS, Chen X, Obermeier P, Seeber L, et al. Evaluation of novel second-generation RSV and influenza rapid tests at the point of care. Diagn Microbiol Infect Dis 2015;81(3):171–6.

77. Cohen DM, Kline J, May LS, Harnett GE, Gibson J, Liang SY, et al. Accurate PCR detection of influenza A/B and respiratory syncytial viruses by use of Cepheid Xpert Flu+RSV Xpress assay in point-of-care settings: comparison to Prodesse ProFlu. J Clin Microbiol 2018;56(2):e01237–17.

78. Haigh J, Cutino-Moguel MT, Wilks M, Welch CA, Melzer M. A service evaluation of simultaneous near-patient testing for influenza, respiratory syncytial virus, Clostridium difficile and norovirus in a UK district general hospital. J Hosp Infect 2019;103(4):441–6.

79. Wabe N, Lindeman R, Post JJ, Rawlinson W, Miao M, Westbrook JI, et al. Cepheid Xpert(®) Flu/RSV and Seegene Allplex(™) RP1 show high diagnostic agreement for the detection of influenza A/B and respiratory syncytial viruses in clinical practice. Influenza Other Respir Viruses 2021;15:245–53.

80. Zou X, Chang K, Wang Y, Li M, Zhang W, Wang C, et al. Comparison of the Cepheid Xpert Xpress Flu/RSV assay and commercial real-time PCR for the detection of influenza A and influenza B in a prospective cohort from China. Int J Infect Dis 2019;80:92–7.

81. Ginocchio CC, Swierkosz E, McAdam AJ, Marcon M, Storch GA, Valsamakis A, et al. Multicenter study of clinical performance of the 3M Rapid Detection RSV test. J Clin Microbiol 2010;48(7):2337–43.

82. Munjal I, Gialanella P, Goss C, McKitrick JC, Avner JR, Pan Q, et al. Evaluation of the 3M rapid detection test for respiratory syncytial virus (RSV) in children during the early stages of the 2009 RSV season. J Clin Microbiol 2011;49(3):1151–3.

83. Juretschko S, Mahony J, Buller RS, Manji R, Dunbar S, Walker K, et al. Multicenter clinical evaluation of the Luminex Aries Flu A/B & RSV assay for pediatric and adult respiratory tract specimens. J Clin Microbiol 2017;55(8):2431–8.

84. Huang HS, Tsai CL, Chang J, Hsu TC, Lin S, Lee CC. Multiplex PCR system for the rapid diagnosis of respiratory virus infection: systematic review and meta-analysis. Clin Microbiol Infect 2018;24(10):1055–63.

85. Leber AL, Everhart K, Daly JA, Hopper A, Harrington A, Schreckenberger P, et al. Multicenter evaluation of BioFire FilmArray Respiratory Panel 2 for detection of viruses and bacteria in nasopharyngeal swab samples. J Clin Microbiol 2018;56(6):e01945–17.

86. Vos LM, Riezebos-Brilman A, Schuurman R, Hoepelman AIM, Oosterheert JJ. Syndromic sample-to-result PCR testing for respiratory infections in adult patients. Neth J Med 2018;76(6):286–93.

87. Aslanzadeh J, Zheng X, Li H, Tetreault J, Ratkiewicz I, Meng S, et al. Prospective evaluation of rapid antigen tests for diagnosis of respiratory syncytial virus and human metapneumovirus infections. J Clin Microbiol 2008;46(5):1682–5.

88. Vaz-de-Lima LR, Souza MC, Matsumoto T, Hong MA, Salgado MM, Barbosa ML, et al. Performance of indirect immunofluorescence assay, immunochromatography assay and reverse transcription-polymerase chain reaction for detecting human respiratory syncytial virus in nasopharyngeal aspirate samples. Mem Inst Oswaldo Cruz 2008;103(5):463–7.

89. Goodrich JS, Miller MB. Comparison of Cepheid’s analyte-specific reagents with BD directigen for detection of respiratory syncytial virus. J Clin Microbiol 2007;45(2):604–6.

90. Babady NE, England MR, Jurcic Smith KL, He T, Wijetunge DS, Tang YW, et al. Multicenter evaluation of the ePlex Respiratory Pathogen Panel for the detection of viral and bacterial respiratory tract pathogens in nasopharyngeal swabs. J Clin Microbiol 2018;56(2):e01658–17.

91. Sam SS, Caliendo AM, Ingersoll J, Abdul-Ali D, Hill CE, Kraft CS. Evaluation of performance characteristics of Panther Fusion Assays for detection of respiratory viruses from nasopharyngeal and lower respiratory tract specimens. J Clin Microbiol 2018;56(8):e00787–18.

92. Pichon M, Valette M, Schuffenecker I, Billaud G, Lina B. Analytical performances of the Panther Fusion System for the detection of respiratory viruses in the French National Reference Centre of Lyon, France. Microorganisms 2020;8(9):1371.

93. Stellrecht KA, Cimino JL, Wilson LI, Maceira VP, Butt SA. Panther Fusion® respiratory virus assays for the detection of influenza and other respiratory viruses. J Clin Virol 2019;121:104204.

94. Voermans JJC, Mulders D, Pas SD, Koopmans MPG, van der Eijk AA, Molenkamp R. Performance evaluation of the Panther Fusion® respiratory tract panel. J Clin Virol 2020;123:104232.

95. Landry ML, Ferguson D. Comparison of Simplexa Flu A/B & RSV PCR with cytospin-immunofluorescence and laboratory-developed TaqMan PCR in predominantly adult hospitalized patients. J Clin Microbiol 2014;52(8):3057–9.

