## Supplementary materials for "Diagnostic Accuracy of Commercially Available Tests for Respiratory Syncytial Virus: A Scoping Literature Review in the COVID-19 Era"

| **Supplemental Table 1.** **Data sources and search strategy.** | |
| --- | --- |
| **PubMed, search conducted on January 21, 2021** | |
| **Search number** | **Query** |
| #1 | "Respiratory syncytial virus, Human"[Mesh] OR "Respiratory syncytial virus infections"[Mesh] OR "Bronchiolitis, Viral"[Mesh] OR "respiratory syncytial virus"[tiab] OR bronchiolitis[tiab] OR RSV[tiab] |
| #2 | "Sensitivity"[tiab] OR "Sensitive"[tiab] OR "Specificity"[tiab] OR "Specific"[tiab] OR "Accuracy"[tiab] OR "Accurate"[tiab] |
| #3 | #1 AND #2 |
| #4 | "Immunoassay"[Mesh] OR "Reagent kits, diagnostic"[Mesh] OR "Antigens, viral"[Mesh] OR immunoassay*[tiab] OR immunochromatograph*[tiab] OR "rapid antigen test*"[tiab] OR "rapid antigen detection test*"[tiab] OR "rapid antigen detection"[tiab] OR "antigen detection"[tiab] OR "antigen test*"[tiab] OR "RADT"[tiab] |
| #5 | "Diagnosis"[Majr] OR "Diagnostic Techniques and Procedures"[Majr] OR "Diagnostic Equipment"[Majr] OR "Biological Assay"[Majr] OR "Biological Assay"[Majr] OR "rapid test*"[tiab] OR "rapid detection test*"[tiab] OR "rapid diagnos*"[tiab] OR "rapid detection"[tiab] OR "Quick testing"[tiab] OR "Point of care"[tiab] OR "Point–of–care"[tiab] OR "Bedside testing"[tiab] OR "rapid diagnostic test*"[tiab] |
| #6 | "Nucleic Acids"[Majr] OR "Polymerase Chain Reaction"[Majr] OR "Nucleic acid"[tiab] OR "Nucleic acids"[tiab] OR "PCR"[tiab] OR "Polymerase Chain Reaction"[tiab] OR "Polymerase Chain Reactions"[tiab] OR "RT–PCR"[tiab] OR "Real–time PCR"[tiab] OR "Real time PCR"[tiab] OR "Real–time polymerase chain reaction"[tiab] OR "Real–time polymerase chain reactions"[tiab] OR "Real time polymerase chain reaction"[tiab] OR "Real time polymerase chain reactions"[tiab] OR "Kinetic polymerase chain reaction"[tiab] OR "Kinetic PCR"[tiab] OR "Molecular"[tiab] OR "Molecular test"[tiab] |
| #7 | "Primary Cell Culture"[Majr] OR "Viral culture"[tiab] OR "Tissue culture"[tiab] OR "Culture"[tiab] |
| #8 | #3 AND #4 |
| #9 | #3 AND #5 |
| #10 | #3 AND #6 |
| #11 | #3 AND #7 |
| #12 | #8 OR #9 OR #10 OR #11 |
| #13 | #8 OR #9 OR #10 OR #11 |
| #14 | #8 OR #9 OR #10 OR #11 |
| #15 | #14 NOT ("animals"[Mesh] NOT "humans"[Mesh]) |
| #16 | "Review"[Publication Type] OR "Editorial"[Publication Type] OR "Case Reports" [Publication Type] OR "addresses"[Publication Type] OR "biography"[Publication Type] OR "case reports"[Publication Type] OR "comment"[Publication Type] OR "directory"[Publication Type] OR "festschrift"[Publication Type] OR "interview"[Publication Type] OR "lectures"[Publication Type] OR "legal cases"[Publication Type] OR "legislation"[Publication Type] OR "news"[Publication Type] OR "newspaper article"[Publication Type] OR "patient education handout"[Publication Type] OR "popular works"[Publication Type] OR "case report"[Tiab] OR "report a case"[Tiab] |
| #17 | ((("systematic"[Tiab] OR "systematically"[Tiab]) AND ("review"[Tiab] OR "review"[Tiab] OR "reviewing"[Tiab])) OR "systematic"[sb] OR "Meta–Analysis"[Publication Type] OR "Meta analysis"[Tiab] OR "Meta analyses"[Tiab] OR "Meta–analysis"[Tiab] OR "Meta–analyses"[Tiab] OR "Meta–Analysis as topic"[Mesh]) |
| #18 | #16 NOT #17 |
| #19 | #15 NOT #18 |
| **Embase, search conducted on January 21, 2021** | |
| **Search number** | **Query** |
| #1 | 'human respiratory syncytial virus'/exp OR 'human respiratory syncytial virus' OR 'respiratory syncytial virus infection'/exp OR 'respiratory syncytial virus infection' OR 'viral bronchiolitis'/exp OR 'viral bronchiolitis' OR 'respiratory syncytial virus':ti,ab OR bronchiolitis:ti,ab OR rsv:ti,ab |
| #2 | 'sensitivity':ti,ab OR 'sensitive':ti,ab OR 'specificity':ti,ab OR 'specific':ti,ab OR 'accuracy':ti,ab OR 'accurate':ti,ab |
| #3 | #1 AND #2 |
| #4 | 'immunoassay'/exp OR 'diagnostic kit'/exp OR 'virus antigen'/exp OR immunoassay*:ti,ab OR immunochromatograph*:ti,ab OR 'rapid antigen test*':ti,ab OR 'rapid antigen detection test*':ti,ab OR 'rapid antigen detection':ti,ab OR 'antigen detection':ti,ab OR 'antigen test*':ti,ab OR 'radt':ti,ab |
| #5 | 'diagnosis'/mj OR 'diagnostic procedure'/mj OR 'diagnostic equipment'/mj OR 'bioassay'/mj OR 'rapid test*':ti,ab OR 'rapid detection test*':ti,ab OR 'rapid diagnos*':ti,ab OR 'rapid detection':ti,ab OR 'quick testing':ti,ab OR 'point of care':ti,ab OR 'point–of–care':ti,ab OR 'bedside testing':ti,ab OR 'rapid diagnostic test*':ti,ab |
| #6 | 'nucleic acids, nucleic acid components and their derivatives'/mj OR 'polymerase chain reaction'/mj OR 'nucleic acid':ti,ab OR 'nucleic acids':ti,ab OR 'pcr':ti,ab OR 'polymerase chain reaction':ti,ab OR 'polymerase chain reactions':ti,ab OR 'rt–pcr':ti,ab OR 'real–time pcr':ti,ab OR 'real time pcr':ti,ab OR 'real–time polymerase chain reaction':ti,ab OR 'real–time polymerase chain reactions':ti,ab OR 'real time polymerase chain reaction':ti,ab OR 'real time polymerase chain reactions':ti,ab OR 'kinetic polymerase chain reaction':ti,ab OR 'kinetic pcr':ti,ab OR 'molecular':ti,ab OR 'molecular test':ti,ab |
| #7 | 'primary cell culture'/mj OR 'viral culture':ti,ab OR 'tissue culture':ti,ab OR 'culture':ti,ab |
| #8 | #3 AND #4 |
| #9 | #3 AND #5 |
| #10 | #3 AND #6 |
| #11 | #3 AND #7 |
| #12 | #8 OR #9 OR #10 OR #11 |
| #13 | #12 AND [english]/lim AND [2005–2021]/py |
| #14 | #13 NOT ('animal'/exp NOT 'human'/exp) |
| #15 | #14 AND ('article'/it OR 'article in press'/it OR 'chapter'/it OR 'editorial'/it OR 'erratum'/it OR 'letter'/it OR 'note'/it OR 'review'/it OR 'short survey'/it) |
| #16 | #14 AND ('conference abstract'/it OR 'conference paper'/it) |
| #17 | 'review':it OR 'editorial':it OR 'addresses':it OR 'biography':it OR 'case reports':it OR 'comment':it OR 'directory':it OR 'festschrift':it OR 'interview':it OR 'lectures':it OR 'legal cases':it OR 'legislation':it OR 'news':it OR 'newspaper article':it OR 'patient education handout':it OR 'popular works':it OR 'case report':ti,ab OR 'report a case':ti,ab |
| #18 | ('systematic':ti,ab OR 'systematically':ti,ab) AND ('review':ti,ab OR 'reviewing':ti,ab) OR 'systematic':it OR 'meta–analysis':it OR 'meta analyses':ti,ab OR 'meta–analysis':ti,ab OR 'meta–analyses':ti,ab OR 'meta analysis (topic)'/exp |
| #19 | #17 NOT #16 |
| #20 | #15 NOT #19 |

| **Supplemental Table 2.** **Summary of all commercially available RSV tests included in the literature review.** | | | | | | |
| --- | --- | --- | --- | --- | --- | --- |
| **Assay name** | **Manufacturer** | **CLIA-waived status*^a^*** | **Methodology** | **Technology** | **Virus(es) detected** | **Analytical time**  **(min)** |
| **RSV tests with analytical time ≤30 min** | | | | | | |
| 3M Rapid Detection RSV Test | 3M Health Care | Moderate | Lateral flow chromatographic immunoassay | Antigen | RSV only | 15 |
| Alere BinaxNOW RSV | Abbott | Waived | Lateral flow chromatographic immunoassay | Antigen | RSV only | 15 |
| BD Veritor System RSV | Becton, Dickinson and Company | Waived | Lateral flow chromatographic immunoassay | Antigen | RSV only | 10 |
| Bioline RSV | Abbott | No | Chromatographic immunoassay | Antigen | RSV only | 15 |
| cobas influenza A/B & RSV Assay | Roche Diagnostics | Waived | RT-PCR | Molecular | RSV/Influenza | 20 |
| Colloidal Gold Genesis | Genesis Company | No | Rapid antigen detection test | Antigen | RSV only | Not reported |
| Directigen EZ RSV | Becton, Dickinson and Company | Moderate | Lateral flow chromatographic immunoassay | Antigen | RSV only | 15 |
| GenRead RSV | Orion Diagnostica Oy | No | Reverse transcription strand invasion-based amplification | Molecular | RSV only | Not reported |
| Humasis RSV Antigen Test | Humasis | No | Lateral flow chromatographic immunoassay | Antigen | RSV only | 15 |
| ID NOW RSV*^b^* | Abbott | Waived | Isothermal nucleic acid amplification assay | Molecular | RSV only | 13 |
| QuickVue RSV | Quidel | Waived | Dipstick chromatographic immunoassay | Antigen | RSV only | 15 |
| RSV K-SeT | Coris BioConcept | No | Membrane technology with colloidal gold nanoparticles | Antigen | RSV only | 15 |
| RSV Respi-Strip | Coris BioConcept | No | Dipstick chromatographic immunoassay | Antigen | RSV only | 15 |
| Simprova-RV | Eiken Chemical | No | Loop-mediated isothermal nucleic acid amplification assay | Molecular | RSV/Influenza/ HMPV | 30 |
| Sofia RSV FIA | Quidel | Waived | Antigen detection based lateral flow  immunoassay | Antigen | RSV only | 15 |
| Thermo Electron RSV OIA kit | Thermo BioStar | No | Optical immunoassay | Antigen | RSV only | <20 |
| TRU RSV | Meridian Bioscience | No | Lateral flow chromatographic immunoassay | Antigen | RSV only | 15 |
| Xpect RSV | Thermo Scientific | Waived | Lateral flow chromatographic immunoassay | Antigen | RSV only | 15 |
| Xpert Flu/RSV XC | Cepheid | Moderate | RT-PCR | Molecular | RSV/Influenza | Not reported |
| Xpert Xpress Flu/RSV | Cepheid | Waived | RT-PCR | Molecular | RSV/Influenza | 30 |
| **RSV tests with analytical time >30 min** | | | | | | |
| Allplex Respiratory Panel 1 | Seegene Inc | No | RT-PCR | Molecular | Multiplex | Not reported |
| Aries Flu A/B & RSV Assay | Luminex Corporation | Moderate | RT-PCR | Molecular | RSV/Influenza | 120 |
| BioFire FilmArray Respiratory 2.1 Panel | bioMérieux | Moderate | RT-PCR | Molecular | Multiplex | 45 |
| CLART PneumoVir | Genomica | No | RT-PCR DNA microarray | Molecular | Multiplex | Not reported |
| ePlex Respiratory Pathogen Panel | GenMark Diagnostics | Moderate | RT-PCR | Molecular | Multiplex | 120 |
| Magicplex RV Panel Real-Time Test | Seegene Inc | No | RT-PCR | Molecular | Multiplex | <5 hours |
| mariPOC Respi test | ArcDia | No | Sandwich immunoassay | Antigen | Multiplex | 20–120*^c^* |
| MultiCode-PLx Respiratory Virus Panel | EraGen Biosciences, Inc. | No | RT-PCR | Molecular | Multiplex | Not reported |
| nCounter | NanoString Technologies | No | Digital method of mRNA expression quantification | Molecular | Multiplex | Not reported |
| NucliSens EasyQ Respiratory Syncytial Virus A+B assay | bioMerieux | No | Nucleic acid sequence-based amplification | Molecular | RSV only | <4 hours |
| NxTAG-Respiratory  Pathogen Panel | Luminex Corporation | High | RT-PCR with bead hybridization | Molecular | Multiplex | Not reported |
| Panther Fusion Flu A/B/RSV Assay | Hologic, Inc. | High | RT-PCR | Molecular | RSV/Influenza | Not reported |
| Prodesse ProFlu+ Assay | Hologic, Inc. | High | RT-PCR | Molecular | RSV/Influenza | 4 hours |
| QIAstat-Dx Respiratory Panel | Qiagen | Moderate | RT-PCR | Molecular | Multiplex | 60 |
| Seeplex  RV15 OneStep ACE Detection | Seegene Inc | No | RT-PCR | Molecular | Multiplex | Not reported |
| Simplexa Flu A/B & RSV | Diasorin Molecular | High | RT-PCR | Molecular | RSV/Influenza | 60 |
| Solana RSV + hMPV | Quidel | Moderate | Reverse Transcriptase-Helicase-Dependent Amplification | Molecular | RSV/ HMPV | 45 |
| Speed-Oligo RSV | Vircell | No | RT-PCR | Molecular | RSV only | 90 |
| Verigene Respiratory Virus Plus Nucleic Acid Test | Nanosphere | Moderate | RT-PCR | Molecular | Multiplex | Not reported |
| *^a^*CLIA status is taken from: <https://www.accessdata.fda.gov/scripts/cdrh/cfdocs/cfClia/Search.cfm>, last accessed 04 October 2021. Information regarding the methodology and analytical time of the tests are taken from the respective manufacturers’ data sheets.  *^b^*The ID NOW RSV was formerly known as the Alere i RSV.  *^c^*On average, the mariPOC Respi test reports 80% of positive samples within 20 min and 90% of positive results within 2 hours. The final result within 2 hours reports low positive and negative results.  CLIA, Clinical Laboratory Improvement Amendments; DNA, deoxyribonucleic acid; HMPV, human metapneumovirus; mRNA, messenger ribonucleic acid; RSV, respiratory syncytial virus; RT-PCR, reverse transcription-polymerase chain reaction. | | | | | | |

| **Supplemental** **Table 3. Sensitivity and specificity of RSV tests by viruses detected in all included studies.** | | | |
| --- | --- | --- | --- |
| **Viruses detected** | **RSV only** | **RSV/Influenza** | **Multiplex** |
| Number of sub-records, n (%) | 70 (52.6) | 27 (20.3) | 33 (24.8)*^a^* |
| **Overall** | | | |
| Sensitivity, % | 25.7–100 | 66.7–100 | 50.0–100 |
| Specificity, % | 77–100 | 94.3–100 | 91.5–100 |
| **Tests using molecular technology only** |  |  |  |
| Sensitivity, % | 93–100 | 66.7–100 | 62.5–100 |
| Specificity, % | 77–100 | 94.3–100 | 91.5–100 |
| *^a^*RSV tests that detected HMPV, Simprova-RV (two sub-records) and Solana RSV + hMPV (one sub-record), were not included in this analysis. HMPV, human metapneumovirus; RSV, respiratory syncytial virus. | | | |

| **Supplemental Table 4.** **Sensitivity and specificity of all commercially available RSV tests included in the literature review.** | | | |
| --- | --- | --- | --- |
| **Test** | **Sensitivity range, %  (95% CI)** | **Specificity range, %  (95% CI)** | **References** |
| **RSV tests with analytical time ≤30 min** | | | |
| 3M Rapid Detection RSV Test | 60.0 (38.5–81.5)  –  87.3 (83.8–90.1) | 95.6 (93.8–96.9)  –  99.6 (98.7–100) | (1, 2) |
| BD Veritor System RSV | 67.5 (56.1–77.6)  –  97.6 (NR) | 96.8 (91.1–99.3)  –  100 (97.0–100) | (3-10) |
| Binax NOW RSV | 41.2 (NR)  –  90 (NR) | 93.2 (92.8–93.6)  –  100.0 (97–100) | (4, 7, 11-21) |
| cobas influenza A/B & RSV Assay | 94.2 (87.9–97.9)  –  100.0 (96.07–100.0) | 94.29 (86.01–98.42)  –  100 (97.7–100) | (22-28) |
| Directigen EZ RSV | 59 (NR)  –  90 (NR) | 89.5 (NR)  –  99.5 (97–100) | (9, 12, 19, 29-31) |
| ID NOW RSV*^a^* | 93 (89–96)  –  100 (93–100) | 96 (93–98)  –  98.0 (95.8–99.1) | (27, 32-34) |
| QuickVue RSV | 25.7 (NR)  –  90.1 (86.8–93.4) | 98.5 (NR)  –  99.5 (92–99) | (9, 35-37) |
| RSV Respi-Strip | 36.8 (16.3–61.6)  –  92 (86–96) | 90.2 (83.9–94.7)  –  99 (97–100) | (12, 38, 39) |
| Sofia RSV FIA | 74.8 (68.0–80.9)  –  100 (82–100) | 86.8 (85.7–87.4)  –  100 (95.2–100) | (4, 8, 9, 18, 20, 40-42) |
| Xpert Flu/RSV XC | 90.6 (NR)  –  100 (80–100) | 99.4 (NR)  –  100 (91.9–100) | (43-45) |
| Xpert Xpress Flu/RSV | 66.7 (24.1–94.0)  –  98.1 (88.8–99.9) | 98.1 (96.6–99.0)  –  100 (99.7–100) | (23, 46-49) |
| **RSV tests with analytical time >30 min** | | | |
| Aries Flu A/B & RSV Assay | 88.6 (58.3–97.6)  –  97.1 (94.4–98.7) | 98.4 (97.7–98.9)  –  100.0 (98.8–100) | (21, 23, 24, 50) |
| BioFire FilmArray Respiratory 2.1 Panel | 62.5 (24.5–91.5)  –  99.4 (96.9–99.9) | 98.3 (97.5–98.9)  –  100 (97.2–100) | (23, 51-53) |
| ePlex Respiratory Pathogen Panel | 89.6 (80.0–94.8)  –  100 (92.6–100) | 98.9 (94.2–99.8)  –  100 (99.8–100) | (54, 55) |
| mariPOC Respi test | 50.0 (22.3–77.7)  –  90 (79.5–96.2) | 98.3 (NR)  –  100 (97.5–100) | (56-58) |
| Panther Fusion Flu A/B/RSV Assay | 88.4 (81.1–93.1)  –  100 (NR) | 98.7 (97.0–99.6)  –  100 (99.7–100) | (23, 24, 55, 59-61) |
| Simplexa Flu A/B & RSV | 73.3 (44.8–91.0)  –  87.0 (74.5–94.2) | 99.4 (96.3–99.9)  –  100 (98.9–100) | (23, 24, 62) |
| Sensitivity and specificity ranges were not reported for RSV tests with <3 supporting sub-records. *^a^*The ID NOW RSV was formerly known as the Alere i RSV.  CI, confidence interval; FIA, fluorescence immunoassay; NR, not reported; RSV, respiratory syncytial virus. | | | |

38.	Gregson D, Lloyd T, Buchan S, Church D. Comparison of the RSV respi-strip with direct fluorescent-antigen detection for diagnosis of respiratory syncytial virus infection in pediatric patients. J Clin Microbiol 2005;43(11):5782-3.

39.	Newman H, Tshabalala D, Mabunda S, Nkosi N, Carelson C. Rapid testing for respiratory syncytial virus in a resource-limited paediatric intensive care setting. Afr J Lab Med 2020;9(1):a1084.

40.	Gomez S, Prieto C, Folgueira L. A prospective study to assess the diagnostic performance of the Sofia(®) Immunoassay for influenza and RSV detection. J Clin Virol 2016;77:1-4.

41.	Tran LC, Tournus C, Dina J, Morello R, Brouard J, Vabret A. SOFIA(®)RSV: prospective laboratory evaluation and implementation of a rapid diagnostic test in a pediatric emergency ward. BMC Infect Dis 2017;17(1):452.

42.	Tuttle R, Weick A, Schwarz WS, Chen X, Obermeier P, Seeber L, et al. Evaluation of novel second-generation RSV and influenza rapid tests at the point of care. Diagn Microbiol Infect Dis 2015;81(3):171-6.

43.	Popowitch EB, Miller MB. Performance characteristics of Xpert Flu/RSV XC Assay. J Clin Microbiol 2015;53(8):2720-1.

44.	Salez N, Nougairede A, Ninove L, Zandotti C, de Lamballerie X, Charrel RN. Prospective and retrospective evaluation of the Cepheid Xpert® Flu/RSV XC assay for rapid detection of influenza A, influenza B, and respiratory syncytial virus. Diagn Microbiol Infect Dis 2015;81(4):256-8.

45.	Zelyas N, Shokoples S, Droogers J, Lundeberg R, Leedell D, Drews SJ. Performance of the Alere™ i Influenza A&B and the Cepheid Xpert® Flu/RSV XC assays. Future Virology 2017;12(6):251-9.

46.	Cohen DM, Kline J, May LS, Harnett GE, Gibson J, Liang SY, et al. Accurate PCR detection of influenza A/B and respiratory syncytial viruses by use of Cepheid Xpert Flu+RSV Xpress assay in point-of-care settings: comparison to Prodesse ProFlu. J Clin Microbiol 2018;56(2):e01237-17.

57.	Sanbonmatsu-Gámez S, Pérez-Ruiz M, Lara-Oya A, Pedrosa-Corral I, Riazzo-Damas C, Navarro-Marí JM. Analytical performance of the automated multianalyte point-of-care mariPOC® for the detection of respiratory viruses. Diagn Microbiol Infect Dis 2015;83(3):252-6.

58.	Tuuminen T, Suomala P, Koskinen JO. Evaluation of the automated multianalyte point-of-care mariPOC® test for the detection of influenza A virus and respiratory syncytial virus. J Med Virol 2013;85(9):1598-601.

59.	Pichon M, Valette M, Schuffenecker I, Billaud G, Lina B. Analytical performances of the Panther Fusion System for the detection of respiratory viruses in the French National Reference Centre of Lyon, France. Microorganisms 2020;8(9):1371.

60.	Stellrecht KA, Cimino JL, Wilson LI, Maceira VP, Butt SA. Panther Fusion® respiratory virus assays for the detection of influenza and other respiratory viruses. J Clin Virol 2019;121:104204.

61.	Voermans JJC, Mulders D, Pas SD, Koopmans MPG, van der Eijk AA, Molenkamp R. Performance evaluation of the Panther Fusion® respiratory tract panel. J Clin Virol 2020;123:104232.

62.	Landry ML, Ferguson D. Comparison of Simplexa Flu A/B & RSV PCR with cytospin-immunofluorescence and laboratory-developed TaqMan PCR in predominantly adult hospitalized patients. J Clin Microbiol 2014;52(8):3057-9.
